# Systematic Review and Meta-Analysis of Retention and Disengagement After Initiation on Antiretroviral Therapy in Low- and Middle-Income Countries After the Introduction of Universal Test and Treat Policies

**DOI:** 10.1101/2024.12.18.24319250

**Authors:** Amy Zheng, Emma M. Kileel, Alana T. Brennan, David B. Flynn, Sydney Rosen, Matthew P. Fox

## Abstract

**Background:** We previously published a systematic review evaluating retention in care after antiretroviral therapy treatment initiation among adults in low- and middle-income countries. We estimated retention at 36 months to be at 74% for studies published from 2008-2013. This review evaluates retention after the implementation of Universal Test and Treat in 2015.

**Methods:** We searched PubMed, ISI Web of Science, Cochrane Database of Systematic Reviews, and EMBASE for studies published between January 1, 2017, to December 31, 2023 and searched conference abstract repositories from AIDS, IAS, and CROI from 2015-2023. Retention for each study was estimated using 1) simple averages and 2) interpolated for missing timepoints through the last reported timepoint. All-cause attrition for all participants who initiated first-line treatment in low- and middle-income countries was estimated. Overall retention rates were estimated using a generalized linear mixed model with a logit distribution using the interpolated data.

**Results:** A total of 65 studies met our criteria for inclusion for the systematic review. Most studies came from Africa with very few from Europe and Asia. Very few studies reported retention past the first 12 months following treatment initiation. Across all studies, we estimated simple average retention with no interpolation of missing timepoints to be 72.6% at 12 months, 73.2%, at 24 months, 59.8% at 36 months, and 49.4% at 48 months. Utilizing a GLMM model, we estimated retention to be 86.0% at 6 months, 80.0% at 12 months, 82.0% at 24 months, 71.0% at 36 months, and 57.0% at 48 months. In comparison, our prior review conducted in 2015, estimated retention rates to be 91.0% at 6 months, 86.0% at 12 months, 79.0% at 24 months, 75% at 36 months, and 69% at 48 months. These results generally reflect retention at the initiating facility and omit the effect of unreported transfers.

**Conclusions:** Retention in care at 36 months was estimated to be between 60% and 71%. Compared to results from our prior review, retention has stayed largely similar in the post-UTT era. Further research evaluating retention in other geographic areas (i.e., Latin America and the Caribbean, Europe, and Asia) is needed.

**Funding:** ATB and EK was supported by the National Institute of Diabetes and Digestive and Kidney Diseases K01DK116929. SR was supported by INV-031690 from the Gates Foundation to Boston University. AZ was supported by the National Institute of Allergy and Infectious Diseases 1F31AI179292-01A1. Research reported in this publication is supported by the National Institute of Mental Health of the National Institutes of Health under Award Number R01MH121998. The content is solely the responsibility of the authors and does not necessarily represent the official views of the National Institutes of Health. The funders had no role in study design, data collection and analysis, decision to publish, or preparation of the manuscript.

## INTRODUCTION

Despite the dramatic success of HIV treatment programs globally, with nearly 30 million people living with HIV (PLWH) reported to be on antiretroviral therapy (ART), high rates of treatment interruption and disengagement from HIV care persist.^1^ Lifelong retention in HIV care remains a challenge worldwide.^2–5^ Numerous reviews of adults and adolescents in HIV treatment programs have documented varying levels of retention in care over time, as well as a range of barriers to short- and long-term retention.^6–12^ Our own 2015 review found that retention at 36 months following ART initiation averaged between 65 and 70%.^8^ Since retention in care is a prerequisite for viral suppression, which is the ultimate goal of HIV treatment, achieving and sustaining high rates of retention in care is essential.

Since our 2015 review, the landscape of HIV care and treatment has changed dramatically. The most significant development was the adoption of the Universal-Test-and-Treat (UTT) strategy in 2015-2016.^13^ Under UTT, all individuals testing positive for HIV are eligible for immediate ART initiation, rather than waiting for a specific point of disease progression, as in the past.^13,14^ In addition, newer regimens featuring dolutegravir, known for reduced toxicity, have been introduced alongside significant programmatic advancements, including same-day ART initiation and differentiated service delivery (DSD) models for treatment.^13–17^ These changes have been aimed, in part, at improving overall rates of retention in HIV care.^13,14,17–21^

A recent review (2020-2023) focusing solely on sub-Saharan Africa found no difference in retention rates before and after UTT in the first 12 months post ART initiation but did not consider longer time periods or other regions.^7^ Another review found no notable differences in retention between patients enrolled in DSD models compared to patients enrolled in conventional models of care.^10^ Many of these reviews have been limited in scope, looking only at specific care models, drug regimens, regions, or populations.^10,22,23^ Few studies, if any, have explicitly focused on comparing pre- and post-UTT retention to determine whether the policy change—marked by a substantial shift in the overall ART patient population characteristics, including higher average CD4 counts and fewer symptomatic patients—was associated with improvements in retention. As a number of publications have reported retention in routine care among specific cohorts in the years since UTT was adopted, it is an opportune time to conduct an updated review of the literature.^1,13,20^

To evaluate changes in retention on ART in routine care settings since the implementation of UTT, we conducted a systematic review and meta-analysis for low- and middle-income countries (LMICs) globally. Our goals were to: 1) estimate all-cause attrition from ART programs in LMICs in the UTT era; 2) determine how attrition varies across regions; 3) evaluate what proportion of attrition can be attributed to death, transfer out of care, and disengagement from care; and 4) compare these findings to those of our pre-UTT retention review from 2015. We also compare our findings to those of other recent reviews that have considered only specific subsets of the populations we include.

## METHODS

### Search strategy and selection criteria

This systematic review and meta-analysis followed the Preferred Reporting Items for Systematic Reviews and Meta-Analyses (PRISMA) guidelines and is registered with PROSPERO (396758). Ethics review was not required as no human subjects data were used.

We developed our search strategy in consultation with a medical librarian (D.F.). Inclusion and exclusion criteria are listed in Table 1. We limited the review to studies that enrolled participants at ART initiation and provided enough information to estimate the rate of all-cause attrition for at least one of the following time points: 6, 12, 18, 24, 36, 48, 60, 72, 84, and/or 96 months after ART initiation or at the end of the median follow up period. We excluded randomized trials. If observational studies evaluated the impact of an intervention but did not seek to alter retention for the comparison arm, only data from the comparison arm (i.e., those who did not receive the intervention) were included in this review. We included all World Bank defined LMICs.^24^ We made no exclusions based on data quality. When multiple studies reported on a single cohort, we selected the one with the longest follow-up. We accepted each study’s own definitions of attrition and retention, as we did not have access to the primary data to apply a consistent definition. We were unable to distinguish between initiation of ART-naive clients and re-initiation of those returning to care after an interruption, as this characteristic is rarely reported accurately. ^25^

**Table 1.** Study selection inclusion and exclusion criteria.

| **Criterion** | **Inclusion** | **Exclusion** |
| --- | --- | --- |
| Geographic range | Low and middle income countries as defined by the World Bank | High income countries as defined by the World Bank |
| Topic | Treatment of HIV/AIDS with highly active antiretroviral therapy (ART) ($\geq 3$ drug combination, HIV-1 only) | Anything other than treatment of HIV-1 with $\geq 3$-drug ART |
| Language | English | Not English |
| Design | Observational cohort, intention to treat. Must report on outcomes for all participants who discontinued treatment for any reason. | Does not observe a cohort or reports only treatment results or treatment plus mortality results; clinical trial, including Phase 3 trials. |
| Setting | Standard service delivery in any sector (public, nongovernmental, private) that treats the general population. | Research or any other non-standard setting in which national guidelines are not followed. |
| Guidelines in effect | Universal test and treat | Restrictions on treatment eligibility due to CD4 count or disease stage |
| Population | Adults (15+) who initiated or re-initiated first-line ART, including both general populations and special populations such as MSM, prisoners, IDUs, sex workers, transgender persons. | Subsets of general treatment-eligible population (e.g. participants known to have drug resistance or specific baseline CD4 counts). |
| Observation period | Must start at ART initiation or re-initiation, with minimum median follow up of 6 full months (26 weeks); median follow up period must be clearly specified or able to be calculated from information provided; dates of observation period must be reported. | Already on ART at study enrollment. Median follow up of less than 6 full months (<24 weeks) or not specified and not able to be calculated from information provided. |
| Minimum information included | Must report all-cause attrition rate at one of 6, 12, 18, 24, 36, 48, 60, 72, 84 or 96 months or at the end of the median follow up period or provide information needed to calculate one of these rates. | Does not report all-cause attrition rate at at least one of 6, 12, 24, 36, 48, 60, 72, 84 or 96 months or at the end of the median follow up period (and none of these rates can be calculated from information reported) or does not report dates of period of observation. |
| Data Collection | 2015-2023 | Prior to 2015 or after 2023 |

### Search procedures

Two reviewers (A.Z. and E.M.K) independently searched PubMed, ISI Web of Science, Cochrane Database of Systematic Reviews, and EMBASE (full list of search terms in Supplemental Table 1) from January 1, 2017, to December 31, 2023 for eligible publications and abstracts from AIDS (2016, 2018, 2020, 2022), IAS (2015, 2017, 2019, 2021, 2023), and CROI (2015-2023) conferences. Search terms were developed based on the following terms: “attrition,” “engagement,” “disengagement,” “interruption,” “non-adherence,” “retention,” and “loss to follow up”. We conducted a hand search to capture articles not MeSH-indexed and reviewed the reference lists of articles originally selected for data extraction to identify any studies missed.^26^ January 1, 2017 was chosen as the starting date because implementation of UTT began in 2015 and studies published prior to 2017 were likely to report pre-UTT data.

Each citation (title and abstract) was screened in duplicate^27^ for eligibility for full-text review using the criteria in Table 1. Both title/abstract and full text screening were done by two reviewers (AZ/EMK) independently, with any discrepancies resolved by a third reviewer (MPF). Reviewers used a standardized form to extract all relevant information such as definitions of loss to follow-up (LTFU), follow-up time, and percent retained at each timepoint.

Study quality was evaluated using a modified version of the Joanna Briggs Institute (JBI) Critical Appraisal Tool for Prevalence Studies.^28^ The following seven questions were used:: 1) Was the cohort well-defined? (e.g., Question 1 [Was the sample frame appropriate to address the target population] and Question 2 [Were study participants sampled in an appropriate way] from the original tool); 2) Was the definition of loss to follow-up clear and well-defined?; 3) Were loss to follow-up outcomes clearly reported?; 4) Did the study differentiate between “Alive and on ART” and “Retained in Care”?; 5) Did the study differentiate between transfer, death, loss to follow-up, etc.?; 6) Was follow-up time reported, assumed, or calculated? (e.g., Question 8 [Was there appropriate statistical analysis] from the JBI tool); 7) Were valid methods used for the identification of the condition? (e.g., Question 6 unmodified from the JBI tool). Questions 2-5 were based on the following questions from the JBI tool: Question 4: Were the study subjects and the setting described in detail; Question 5: Was the data analysis conducted with sufficient coverage of the identified sample; and Question 6 [Were valid methods used from the identification of the condition. Questions 1-5 had the following response options: “Yes,” “No,” “Unclear,” and “Not Applicable”.^29^ Question 6 had the following responses options: “Reported,” “Assumed,” “Calculated,” or “Not Applicable.” We assigned point values to each response option to calculate an overall score with a maximum score of 11 and a minimum score of −6, with a higher score indicating higher quality. Questions 3 (Was the sample size adequate) and 9 (Was the response rate adequate, and if not, was the low response rate managed appropriately) were not applicable to this quality evaluation as we were evaluating incident outcomes.

### Outcomes and data analysis

To make our results comparable to those of our earlier review, we applied the same methodology, to the extent possible, in this analysis.^8,9^ For each study, we report overall attrition from care at each time point reported in the study and report reasons for attrition, including death, LLTFU, and transfer to another facility. We report median follow-up time for each study. If a study did not report median follow-up, we assumed the median was the last reported timepoint if all participants had the potential to complete the maximum reported follow-up time and retention was ≥50% at end of follow-up. For example, if 80% of participants were retained at 48 months, the median follow-up time would be 48 months. If everyone did not have the potential to complete the maximum reported follow-up, we took the mid-point of the reported minimum and maximum follow-up periods. For example, if a study enrolled participants from 2016 to 2018 and follow-up ended in 2020, some participants would only have the potential for 24 months of follow up while others would have the potential for 48 months. In this scenario, we assumed median follow-up to be 36 months. Two studies reported mean follow-up time and we used that in place of the median.^30,31^

We estimated mean retention at 6, 12, 18, 24, 36, 48, and 60 months in five ways: 1) using simple averages of retention reported by each study; 2) using averages weighted by sample size for each cohort only at time points reported by each study; 3) using weighted averages and interpolated data; 4) using weighted averages and interpolated data excluding any outliers which were defined as an estimate that was greater or less than 2 standard deviations away from the weighted average of the full interpolated dataset and 5) using a generalized linear mixed model (GLMM) with a logit distribution and interpolated data. We interpolated any missing period where possible assuming a linear decline. For example, if a study reported outcomes at 24 months but not at 6 and 12 months, we interpolated 12-month retention as the average of 24-month retention and 100% and interpolated 6-month retention as the average of the interpolated 12-month retention and 100%. We generated bootstrapped 95% confidence intervals utilizing the GLMM estimates with 10,000 samples.^32,33^ We stratified countries into the following regions: Europe and Asia, Latin America and the Caribbean, and Africa to evaluate how retention varied across region.

In our prior reviews, we evaluated retention at each timepoint using a lifetable analysis and Kaplan-Meier curves. In the current review, however, we found that a majority of studies were limited to shorter follow up time periods (e.g., the first 12 months only), unlike in previous reviews. Because of this, a lifetable analysis would result in many study participants being censored and would generate a falsely low estimate of retention in care. We therefore omitted this type of analysis in this review.

To evaluate potential for publication bias, we plotted mean retention by the last time period reported for each study. If studies with longer time periods reported higher retention rates compared to studies with shorter time periods, this could be indicative of publication bias. Because not all studies reported retention for the same time points, we also conducted a sensitivity analysis modelling three scenarios for attrition. The best-case scenario assumed no attrition after the last time point reported up to 72 months. The worst-case scenario assumed attrition continued in the same linear trend as observed. The midpoint scenario was an average of the best-case and worst-case scenarios.

## RESULTS

### Eligible articles and study populations

Our search yielded 11,918 unique articles and abstracts. Of these, 65 met the inclusion criteria (61 articles and 4 abstracts; Figure 1). These studies reported on 65 cohorts and 415,159 individuals (Supplemental Table 2). As in prior reviews, the majority (84.6%) of studies reported on cohorts from sub-Saharan Africa. Of the 55 studies in Africa, 23.6% (19.7% of all studies) were from South Africa. The other ten studies came from the Caribbean (n=1), Asia (n=8), and Europe (n=1). We found no eligible studies from Latin America and were unable to evaluate retention outcomes in this region.

**Figure 1.**
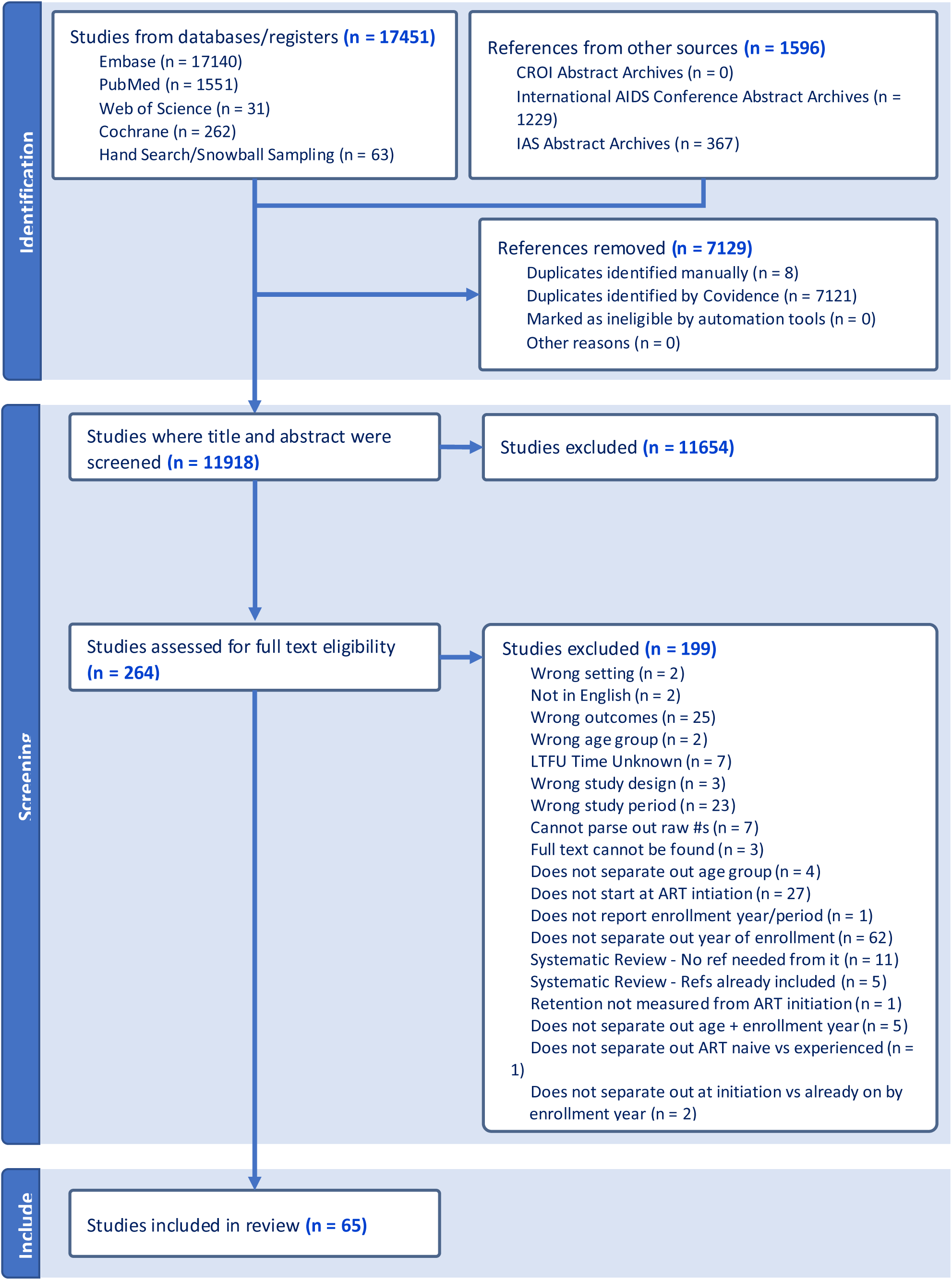
Flowchart of studies included.

Among people initiating ART, 60.4% were female and majority were in their thirties. The median CD4 cell count at initiation ranged from 200-300 cell/mm^3^. People initiating in later years (2017-2019) had slightly higher median CD4 counts than those reported from earlier years (2015-2017). Most studies (89.4%) reported on retention for only the first 12-months after ART initiation (Supplemental Table 2).

Definitions of disengagement from care or loss to follow-up ranged from not returning to the clinic within two months of a scheduled visit (clinic or drug refill) to no return nine months after the initial visit. The most common definition of disengagement was being late for the last scheduled visit or medication pickup by more than 90 days. For studies that made a distinction between LTFU and treatment stoppage, the most common definition for treatment stoppage was the patient was known to be alive but had stopped ART treatment for any reason. For the three studies that reported on treatment interruptions, Dorward et al. defined treatment interruption as not having a visit for 180 days but then having a visit and restarting treatment;^31^ Tlhajoane et al. defined interruptions as returning to care after an absence of >90 days;^34^ and Ibiloye et al. reported the number of patients who experienced a treatment interruption but did not provide a definition.^35^ The range of definitions used in the included studies are presented in Supplemental Table 3.

### Retention outcomes

Table 2 presents the unweighted retention at each time point for each study, weighted averages for overall retention, and a weighted average utilizing interpolated data. Across all studies, simple average retention with no interpolation averaged 72.6% at 12 months, 73.2% at 24 months, 59.8% at 36 months, and 49.4% at 48 months, with substantial heterogeneity across individual studies and countries (Tables 2 and 3). Few studies reported outcomes beyond 36 months (n=5). Results from the weighted average using interpolated data and when excluding outliers were similar and trends were consistent.

**Table 2.** Retention of participants at 6, 12, 18, 24, 36, 48, 60, and 72 months after ART initiation.

| Reference | N | Percentage of participants retained at month: |  |  |  |  |  |  |  |  |  |
| --- | --- | --- | --- | --- | --- | --- | --- | --- | --- | --- | --- |
|  |  | 6 | 12 | 18 | 24 | 36 | 48 | 60 | 72 | 84 | 96 |
| Ahmed 2020<sup>65</sup> | 988 | 86.1 | 79.3 | - | - | - | - | - | - | - | - |
| Alhaj 2019<sup>c 66</sup> | 1492 | - | 74.7 | - | - | - | - | - | - | - | - |
| Atuhaire 2022<sup>67</sup> | 275 | 85.1 | 78.9 | - | 75.3 | - | - | - | - | - | - |
| Avalos 2019<sup>68</sup> | 1523 | - | 95.9 | - | - | - | - | - | - | - | - |
| Benzekri 2022<sup>69</sup> | 207 | - | 59.9 | - | - | - | - | - | - | - | - |
| Byamukama 2022<sup>30</sup> | 976 | - | - | - | - | - | - | 89.6 | - | - | - |
| Cassidy 2023<sup>70</sup> | 1553 | 74.0 | 66.0 | - | - | - | - | - | - | - | - |
| Chauke 2020<sup>71</sup> | 367 | 86.1 | 59.6 | - | - | - | - | - | - | - | - |
| Dorward 2020<sup>31</sup> | 4952 | - | 68.3 | - | - | - | - | - | - | - | - |
| Dumchev 2022<sup>72</sup> | 1057 | - | 90.5 | - | - | - | - | - | - | - | - |
| Eamsakulrat 2022<sup>73</sup> | 270 | - | 84.1 | - | - | - | - | - | - | - | - |
| Ntamatungiro 2021<sup>74</sup> | 902 | - | 59.6 | - | - | - | - | - | - | - | - |
| Eshiwani 2018<sup>75</sup> | 167 | 72.5 | - | - | - | - | - | - | - | - | - |
| Harooni 2022<sup>76</sup> | 124 | - | 94.3 | - | - | - | - | - | - | - | - |
| Hirasen 2020<sup>77</sup> | 1143 | - | 57.3 | - | - | - | - | - | - | - | - |
| Hoang 2022<sup>78</sup> | 4892 | - | 92.2 | - | - | - | - | - | - | - | - |
| Ibiloye 2018<sup>35</sup> | 710 | 63.9 | - | - | - | - | - | - | - | - | - |
| Ibiloye 2021<sup>79</sup> | 3495 | - | 63.5 | - | 55.4 | 51.2 | 46.7 | - | - | - | - |
| Izudi 2022<sup>80</sup> | 9952 | - | 19.1 | - | - | - | - | - | - | - | - |
| Jamieson 2021<sup>81</sup> | 32197 | - | - | 95.7 | - | - | - | - | - | - | - |
| Januraga 2018<sup>82</sup> | 606 | 75.4 | - | - | - | - | - | - | - | - | - |
| Johansson 2021<sup>83</sup> | 2043 | - | 69.6 | - | - | - | - | - | - | - | - |
| Kimanga 2022<sup>84</sup> | 8592 | - | 76.7 | - | - | - | - | - | - | - | - |
| Lilian 2020<sup>85</sup> | 32290 | 69.7 | - | - | - | - | - | - | - | - | - |
| Makurumidze 2020<sup>86</sup> | 3636 | - | - | - | 91.8 | - | - | - | - | - | - |
| Matare 2020<sup>87</sup> | 3971 | 76.7 | 72.3 | - | - | - | - | - | - | - | - |
| Matthews 2020<sup>88</sup> | 437 | - | 94.5 | - | - | - | - | - | - | - | - |
| Mayasi 2022<sup>89</sup> | 11281 | 89.3 | 80.7 | - | 44.0 | 15.3 | 5.4 | - | - | - | - |
| Mshweshwe-Pakela 2020<sup>90</sup> | 710 | 67.2 | - | - | - | - | - | - | - | - | - |
| Nshimirimana 2022<sup>91</sup> | 29829 | - | 97.7 | - | 93.5 | 87.3 | 81.0 | 75.9 | 74.7 | - | - |
| Onoya 2021<sup>92</sup> | 297 | - | 74.4 | - | - | - | - | - | - | - | - |
| Opio 2019<sup>93</sup> | 646 | - | 55.6 | - | - | - | - | - | - | - | - |
| Opito 2020<sup>d 94</sup> | 536 | - | 78.7 | - | - | - | - | - | - | - | - |
| Pillay 2019<sup>95</sup> | 120 | 69.0 | - | - | - | - | - | - | - | - | - |
| Rogers 2021<sup>96</sup> | 423 | - | - | - | 73.0 | - | - | - | - | - | - |
| Romo 2022<sup>97</sup> | 3563 | - | 71.9 | - | - | - | - | - | - | - | - |
| Ross 2019<sup>98</sup> | 1082 | 51.7 | - | - | - | - | - | - | - | - | - |
| Seekaew 2019<sup>99</sup> | 180 | 90.0 | 92.9 | - | - | - | - | - | - | - | - |
| Seekaew 2021<sup>100</sup> | 1904 | 75.0 | 75.1 | - | - | - | - | - | - | - | - |
| Ssempijja 2020<sup>101</sup> | 1305 | - | 84.0 | - | - | - | - | - | - | - | - |
| Stafford 2019<sup>102</sup> | 2652 | - | 59.5 | - | - | - | - | - | - | - | - |
| Stockton 2021<sup>103</sup> | 1091 | 53.2 | - | - | - | - | - | - | - | - | - |
| Teshale 2020<sup>104</sup> | 531 | - | - | - | - | - | 64.6 | - | - | - | - |
| Bernard Marc 2023<sup>105</sup> | 193 | - | 91.7 | - | - | - | - | - | - | - | - |
| Chagomerana 2023<sup>106</sup> | 291 | - | - | - | - | 85.3 | - | - | - | - | - |
| Dorward 2023<sup>107</sup> | 22821 | - | 63.6 | - | - | - | - | - | - | - | - |
| Gemechu 2023<sup>108</sup> | 235 | 68.5 | - | - | - | - | - | - | - | - | - |
|  |  | 6 | 12 | 18 | 24 | 36 | 48 | 60 | 72 | 84 | 96 |
| Govere 2023<sup>109</sup> | 403 | 66.3 | - | - | - | - | - | - | - | - | - |
| Hamooya 2023<sup>110</sup> | 3649 | 91.6 | 92.1 | - | 92.1 | - | - | - | - | - | - |
| Joaquim 2023<sup>111</sup> | 1247 | - | 75.1 | - | - | - | - | - | - | - | - |
| Kimanga 2023<sup>112</sup> | 7046 | 69.0 | - | - | - | - | - | - | - | - | - |
| Lavoie 2023<sup>113</sup> | 75348 | - | 58.0 | - | - | - | - | - | - | - | - |
| MacKellar 2022<sup>114</sup> | 769 | 40.1 | 47.1 | 50.5 | - | - | - | - | - | - | - |
| Masaba 2023<sup>115</sup> | 1515 | 88.4 | 89.0 | - | - | - | - | - | - | - | - |
| Masuke 2023<sup>116</sup> | 9025 | - | 90.0 | - | - | - | - | - | - | - | - |
| Mody 2021<sup>117</sup> | 65673 | - | 59.9 | - | - | - | - | - | - | - | - |
| Mugenyi 2022<sup>19</sup> | 20171 | 77.0 | - | - | - | - | - | - | - | - | - |
| Mwamuye 2022<sup>118</sup> | 786 | 76.5 | 71.0 | 65.6 | 60.8 | - | - | - | - | - | - |
| Nimwesiga 2023<sup>119</sup> | 80 | - | - | 35.0 | - | - | - | - | - | - | - |
| Ojiambo 2023<sup>120</sup> | 328 | 43.3 | 35.7 | - | - | - | - | - | - | - | - |
| Ross 2023<sup>121</sup> | 29017 | - | 64.3 | - | - | - | - | - | - | - | - |
| Singh 2023<sup>122</sup> | 135 | 91.9 | - | - | - | - | - | - | - | - | - |
| Tlhajoane 2021<sup>34</sup> | 829 | - | 47.9 | - | - | - | - | - | - | - | - |
| Zhong 2023<sup>123</sup> | 285 | - | 83.9 | - | - | - | - | - | - | - | - |
| Simple average |  | 73.8 | 72.6 | 61.7 | 73.2 | 59.8 | 49.4 | 82.8 | 74.7 | - | - |
| Weighted average<sup>a,b,f</sup> |  | 75.0 | 66.9 | 93.8 | 79.6 | 66.4 | 59.3 | 76.3 | 74.7 | - | - |
| Weighted average interpolated<sup>a</sup> |  | 82.6 | 70.1 | 88.8 | 80.0 | 67.0 | 59.9 | 76.3<sup>e</sup> | 74.7<sup>e</sup> | - | - |
| Weighted average interpolated with Mayasi 2022 removed<sup>a,g</sup> |  | 82.4 | 69.8 | 92.7 | 89.2 | 83.7 | 77.6 | 76.3<sup>e</sup> | 74.7<sup>e</sup> | - | - |

“-” indicates that these results could not be determined from the report
<sup>a</sup>Weighted by cohort size
<sup>b</sup>Weighted based on reported data
<sup>c</sup>Includes 20 participants who are 10-19 years of age
<sup>d</sup>Includes 37 participants who are < 20 years of age
<sup>e</sup>Fewer studies report to later time points therefore pooled estimates at these timepoints can increase despite using interpolated data
<sup>f</sup>The weighted average for retention at 18-months is much larger at 95.8% because only four studies reported 18-month outcomes and one (Jamieson 2021<sup>81</sup>) of the four had a substantially larger sample size than the other three studies giving it the greatest weight in this calculation
<sup>g</sup>Estimates reported at later timepoints by Mayasi 2022 were exceedingly low and are removed as a sensitivity analysis

**Table 3.**
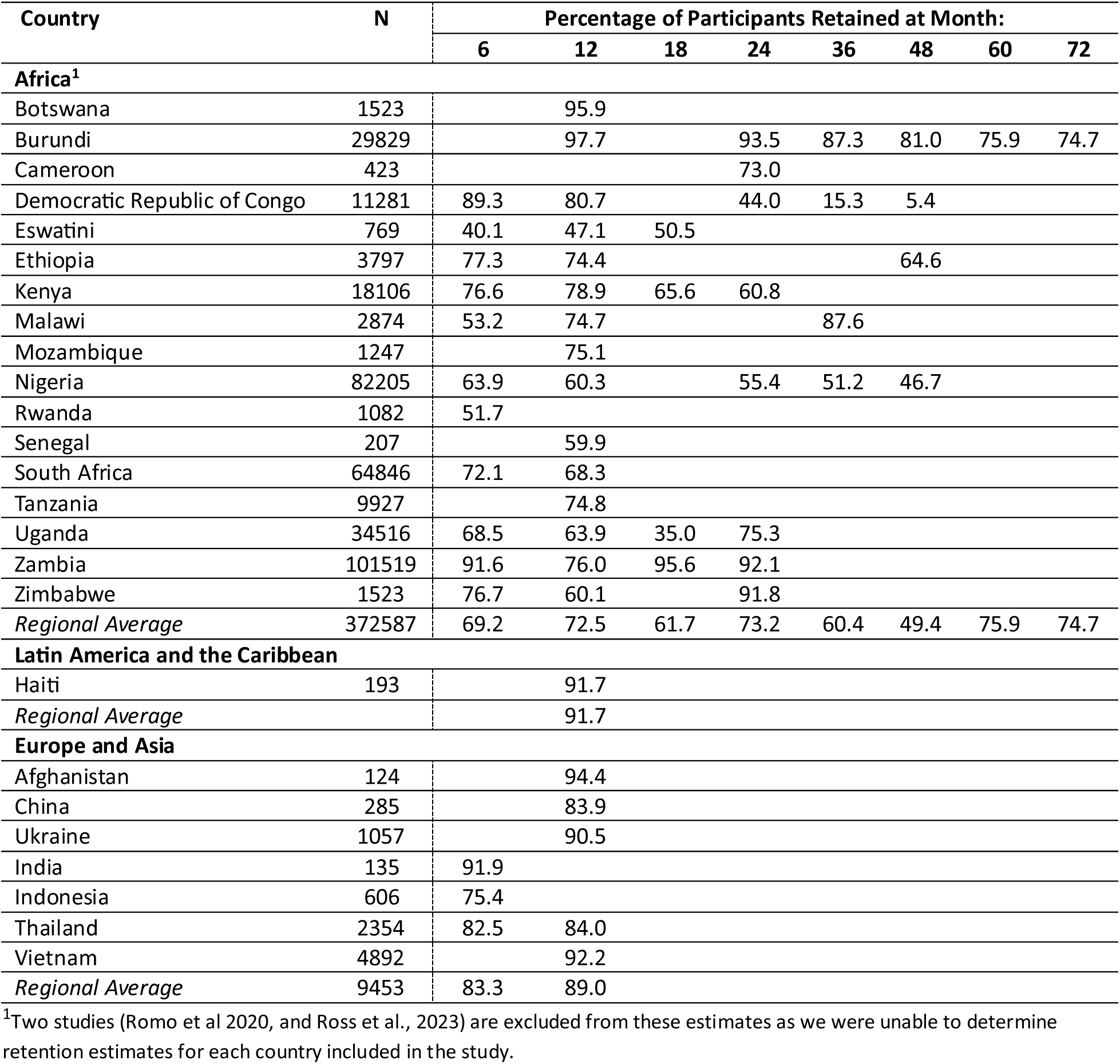
Retention of participants 6, 12, 18, 24, 36, 48, 60, and 72 months after ART initiation by country.

Table 3 presents simple average retention rates by country utilizing only the reported data (i.e., no interpolation). In Figure 2, we illustrate retention rates and their corresponding 95% CIs at each time point up to 36 months, utilizing interpolated data for each study, as well as the pooled estimate from the GLMM model (Supplemental Figure 1 provides estimates at 18, 48, and 60 months). The pooled estimates from the GLMM model were 86.0% at 6 months, 80.0% at 12 months, 82.0% at 24 months, and 71.0% at 36 months. Compared to our 2015 retention review, retention was slightly lower during the UTT period compared to the pre-UTT era (Figure 3).

**Figure 2.**
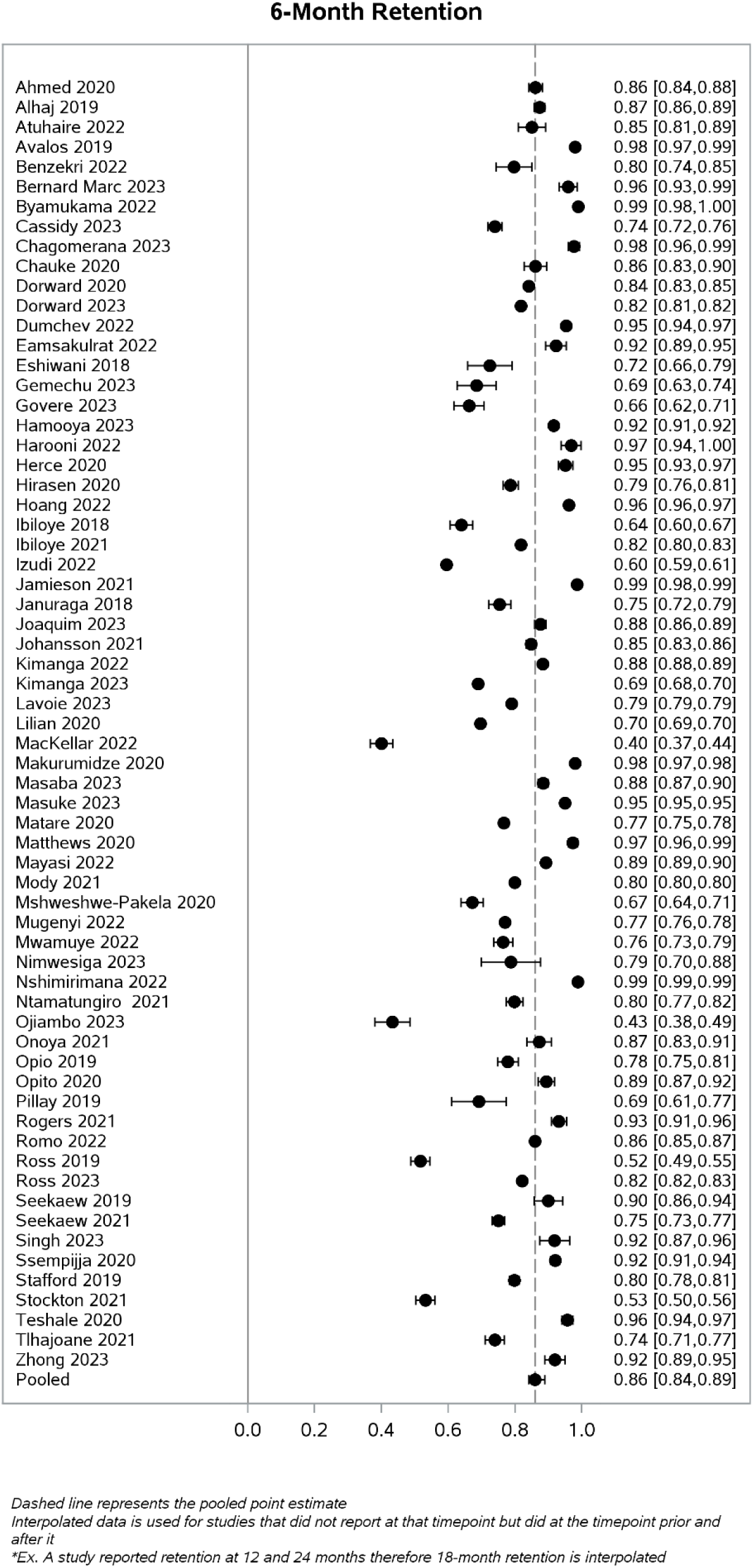
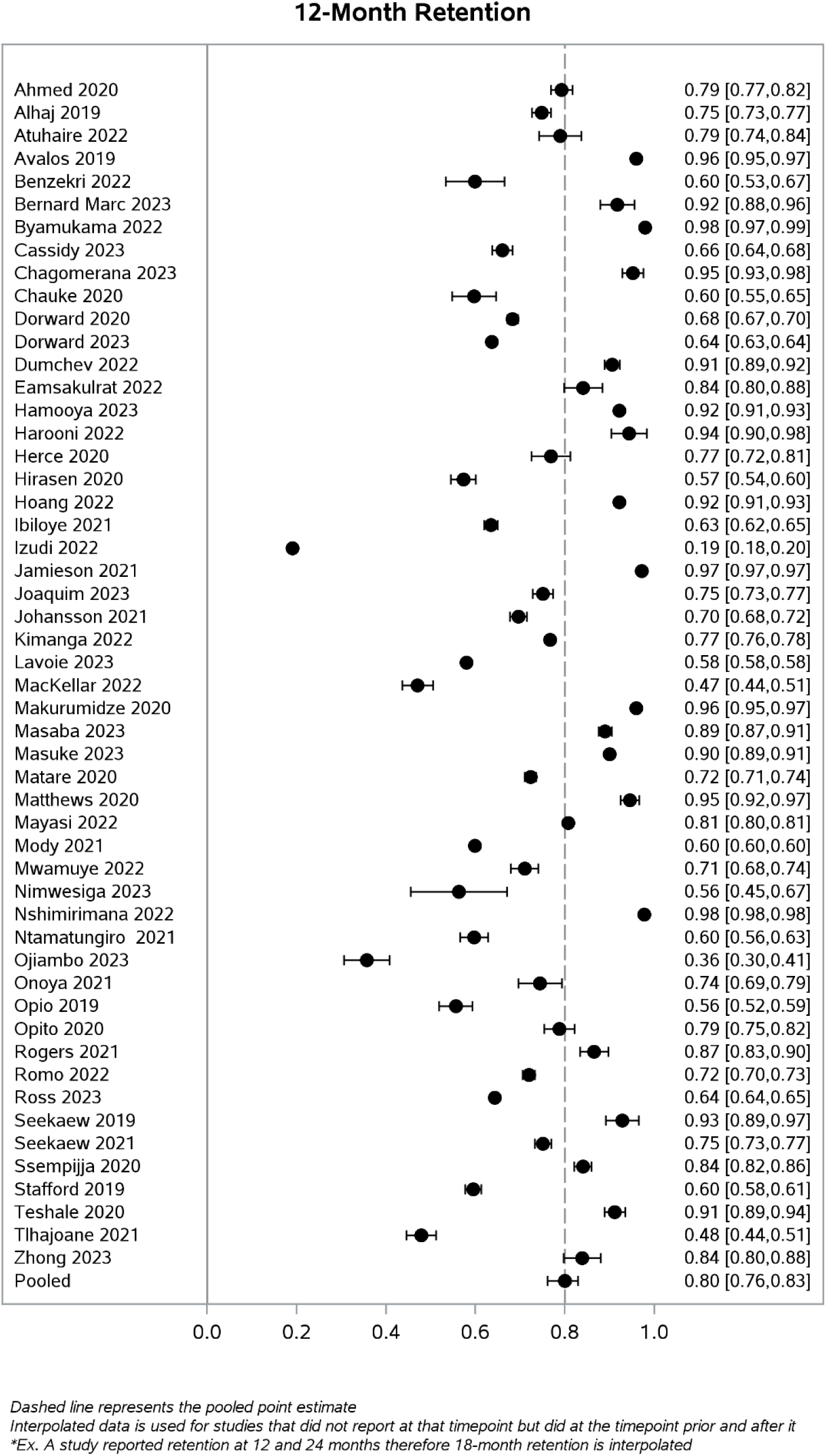
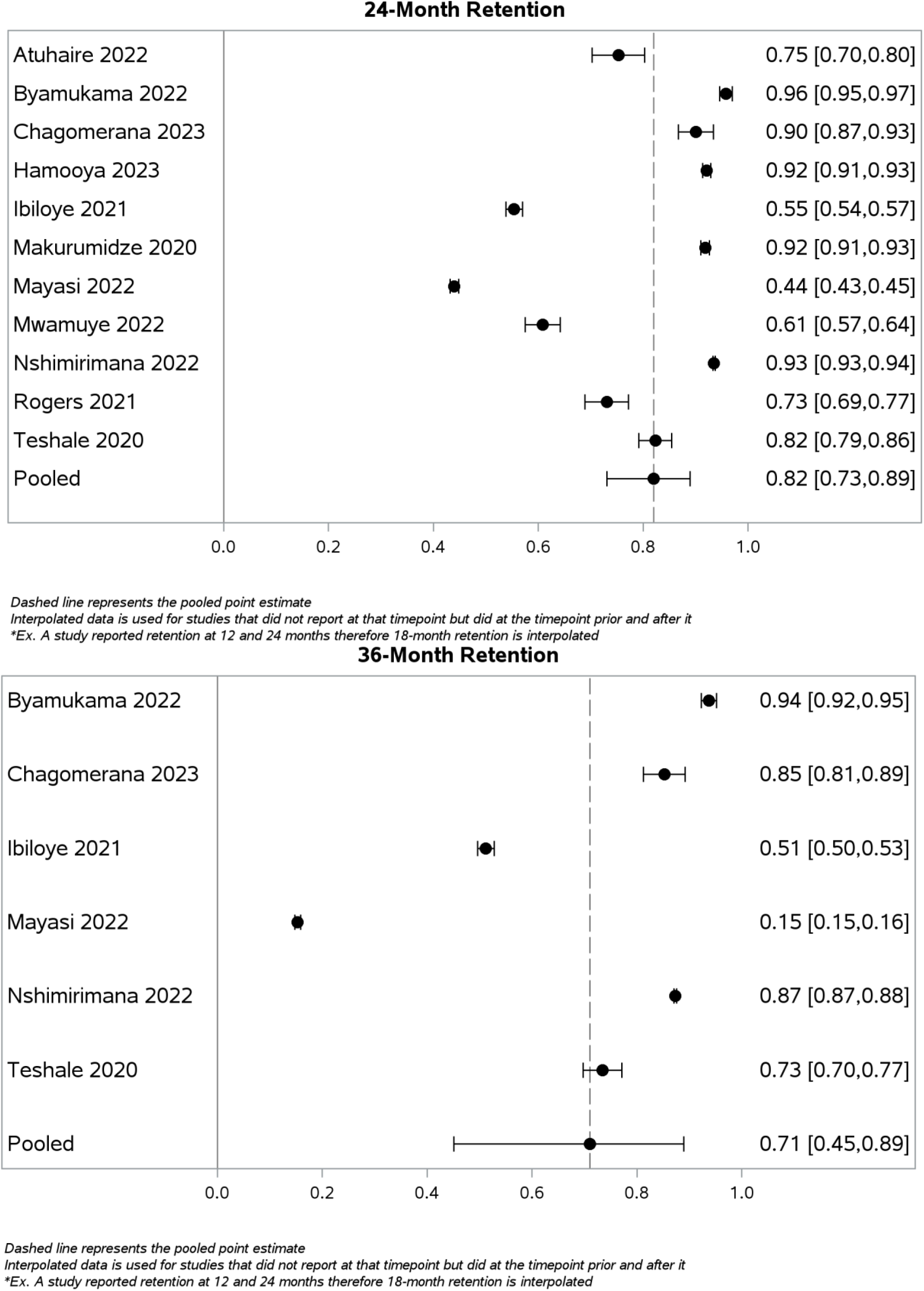
Forest plots of retention rates and their corresponding 95% CIs by time point reported to at 6, 12-, 24- and 36-months.

**Figure 3.**
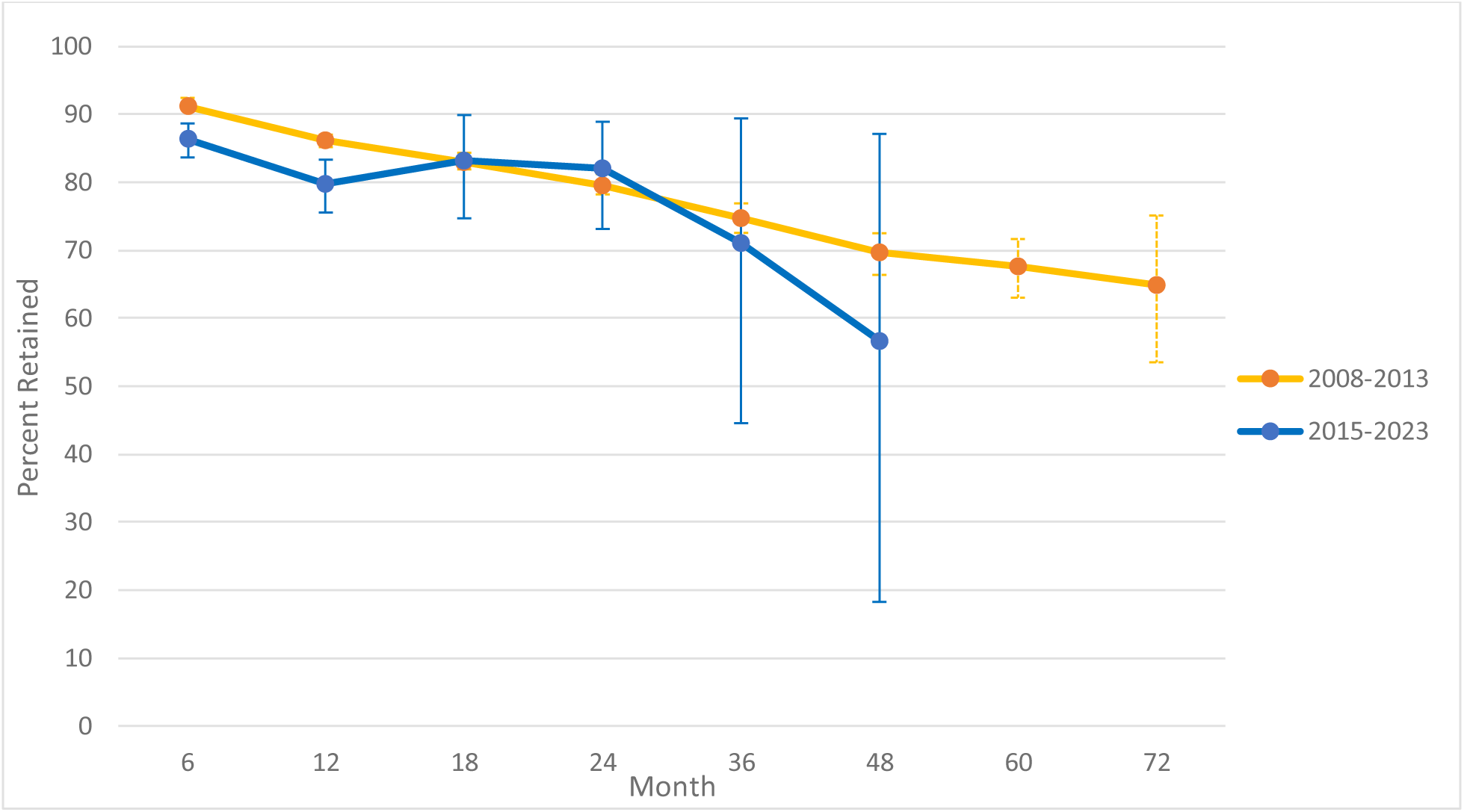
Pooled retention estimates comparing pre-UTT (2008-2013) to UTT (2015-2023) era. ^1^Estimates at 60- and 72-months are not reported for the UTT era given the very few studies that reported data to these timepoints

Reasons for attrition (death, loss to follow-up, transfer) were reported in 60 studies. Table 4 shows overall attrition for each study by reason for attrition. An unweighted average of 2.9% of all participants who initiated ART were known to have died and 22.1% had disengaged from care by the end of the reported follow-up period. Among the 9 that reported treatment stoppage, an average 2.1% of patients stopped treatment. Among the 3 that reported on interruptions, an average of 8.7% of patients reported any treatment interruption.

**Table 4.** Median follow-up and rates of participant attrition, as reported, from antiretroviral treatment programs.

| Reference | Median Follow-Up (Months) | Died (A) | Lost to Follow-Up (B) | Total Attrition from ART (C)<sup>a</sup> | Total Retained (D) | Transferred Care (E) | Total Retained at Original Site (F) |
| --- | --- | --- | --- | --- | --- | --- | --- |
| Ahmed 2020<sup>65</sup> | 12.0 | 5.7 | 15.1 | 20.8 | 79.2 | - | 79.2 |
| Alhaj 2019<sup>b 66</sup> | 12.5 | 3.2 | 14.5 | 25.1 | 74.9 | 7.4 | 67.5 |
| Atuhaire 2022<sup>67</sup> | 24.0 | 0.4 | 26.5 | 26.5 | 73.5 | 10.5 | 63.0 |
| Avalos 2019<sup>68</sup> | 12.0 | 1.9 | 2.2 | 4.1 | 95.9 | - | 95.9 |
| Benzekri 2022<sup>69</sup> | 12.0 | 14.5 | 15.9 | 34.3 | 65.7 | 3.9 | 61.8 |
| Byamukama 2022<sup>c 30</sup> | 34.8 | 2.9 | 2.4 | 10.4 | 89.6 | 5.1 | 84.5 |
| Cassidy 2023<sup>70</sup> | 9.0 | - | 34 | 34.0 | 66.0 | - | 66.0 |
| Chauke 2020<sup>71</sup> | 12.0 | - | 28.4 | 40.4 | 59.6 | 12 | 47.6 |
| Dorward 2020<sup>c 31</sup> | 9.2 | 0.5 | 20.4 | 27.0 | 73.0 | 6.1 | 66.9 |
| Dumchev 2022<sup>72</sup> | 12.0 | - | - | 9.5 | 90.5 | - | 90.5 |
| Eamsakulrat 2022<sup>73</sup> | 12.0 | - | 7.8 | 13.0 | 87.0 | 5.2 | 81.8 |
| Ntamatungiro 2021<sup>74</sup> | 12.0 | 3.7 | 27.7 | 40.1 | 59.9 | 8.8 | 51.1 |
| Eshiwani 2018<sup>75</sup> | 6.0 | 0 | 27.5 | 27.5 | 72.5 | 0 | 72.5 |
| Harooni 2022<sup>76</sup> | 12.0 | - | 5.6 | 5.6 | 94.4 | - | 94.4 |
| Hirasen 2020<sup>77</sup> | 12.0 | 2.6 | 17.2 | 42.6 | 57.4 | 22.8 | 34.6 |
| Hoang 2022<sup>78</sup> | 12.0 | - | 7.8 | 7.8 | 92.2 | - | 92.2 |
| Ibiloye 2018<sup>35</sup> | 7.4 | 3.4 | 23.4 | 36.1 | 63.9 | 9.3 | 54.6 |
| Ibiloye 2021<sup>79</sup> | 42.0 | 1.2 | 46 | 47.2 | 52.8 | - | 52.8 |
| Izudi 2022<sup>80</sup> | 12.0 | 3.3 | 77.6 | 80.9 | 19.1 | - | 19.1 |
| Jamieson 2021<sup>81</sup> | 18.3 | 0.13 | 4.2 | 4.3 | 95.7 | - | 95.7 |
| Januraga 2018<sup>82</sup> | 6.0 | 1.5 | 24.3 | 32.1 | 67.9 | 6.3 | 61.6 |
| Johansson 2021<sup>83</sup> | 12.0 | 6.3 | 24.1 | 30.4 | 69.6 | - | 69.6 |
| Kimanga 2022<sup>84</sup> | 12.0 | 3.1 | 20.3 | 23.3 | 76.7 | - | 76.7 |
| Lilian 2020<sup>85</sup> | 6.0 | 0.6 | 24.2 | 30.3 | 69.7 | 5.5 | 64.2 |
| Makurumidze 2020<sup>d 86</sup> | 17.7 | - | - | 8.2 | 91.8 | - | 91.8 |
| Matare 2020<sup>87</sup> | 12.0 | 0.08 | 27.5 | 27.6 | 72.4 | - | 72.4 |
| Matthews 2020<sup>88</sup> | 8.2 | 0.9 | 4.6 | 5.5 | 94.5 | - | 94.5 |
| Mayasi 2022<sup>89</sup> | 25.0 | - | - | 24.1 | 75.9 | - | 75.9 |
| Mshweshwe-Pakela 2020<sup>90</sup> | 6.0 | - | 32.8 | 32.8 | 67.2 | - | 67.2 |
| Nshimirimana 2022<sup>91</sup> | 66.0 | 3.9 | 25.3 | 25.3 | 74.7 | 8.3 | 66.4 |
| Onoya 2021<sup>92</sup> | 12.0 | - | 34.4 | 34.4 | 65.6 | - | 65.6 |
| Opio 2019<sup>93</sup> | * | 2.5 | 33.4 | 44.4 | 55.6 | 8.5 | 47.1 |
| Opito 2020<sup>e 94</sup> | 12.0 | 1.3 | 12.7 | 21.3 | 78.7 | 7.3 | 71.4 |
| Pillay 2019<sup>95</sup> | 6.0 | - | 31 | 31.0 | 69.0 | - | 69.0 |
| Rogers 2021<sup>96</sup> | 24.0 | 2.6 | 17.3 | 27.0 | 73.0 | 7.1 | 65.9 |
| Romo 2022<sup>97</sup> | 12.0 | 7.9 | 15.2 | 23.2 | 76.8 | - | 76.8 |
| Ross 2019<sup>98</sup> | 9.0 | 1 | 42.7 | 48.3 | 51.7 | 4.6 | 47.1 |
| Seekaew 2019<sup>99</sup> | 9.0 | - | 7.1 | 7.1 | 92.9 | - | 92.9 |
| Seekaew 2021<sup>100</sup> | 9.0 | 0.4 | 24.1 | 24.5 | 75.5 | - | 75.5 |
| Ssempijja 2020<sup>101</sup> | 12.0 | 2 | 10 | 16.0 | 84.0 | 4 | 80.0 |
| Stafford 2019<sup>102</sup> | 12.0 | 2 | 34 | 41.0 | 59.0 | 5 | 54.0 |
| Stockton 2021<sup>103</sup> | 6.0 | 0.2 | - | 6.8 | 93.2 | 6.6 | 86.6 |
| Teshale 2020<sup>104</sup> | 19.6 | 9.6 | 19.21 | 35.4 | 64.6 | 6.6 | 58.0 |
| Bernard Marc 2023<sup>105</sup> | 12.0 | 1.6 | 6.7 | 8.3 | 91.7 | - | 91.7 |
| Chagomerana 2023<sup>106</sup> | 36.0 | 2.3 | 5 | 12.0 | 88.0 | 4.7 | 83.3 |
| Dorward 2023<sup>107</sup> | 12.0 | 1.1 | 21.1 | 36.4 | 63.6 | 14.2 | 49.4 |
| Gemechu 2023<sup>108</sup> | 6.0 | 5.9 | 16.6 | 31.5 | 68.5 | 8.9 | 59.6 |
| Govere 2023<sup>109</sup> | 6.0 | 4.22 | 28.04 | 33.8 | 66.3 | 1.5 | 64.8 |
| Hamooya 2023<sup>110</sup> | 24.0 | 0.2 | 5.9 | 9.3 | 90.7 | 3.2 | 87.5 |
| Joaquim 2023<sup>111</sup> | 12.0 | - | 24.9 | 24.9 | 75.1 | - | 75.1 |
| Kimanga 2023<sup>112</sup> | 6.0 | 1.7 | 24.5 | 31.0 | 69.0 | 4.8 | 64.2 |
| Lavoie 2023<sup>113</sup> | 12.0 | 1.5 | 35.8 | 42.0 | 58.0 | 4.8 | 53.2 |
| MacKellar 2022<sup>114</sup> | 31.6 | 1.8 | 43 | 54.6 | 45.4 | 9.8 | 35.6 |
| Masaba 2023<sup>115</sup> | 11.9 | 0.4 | 1.8 | 3.3 | 96.7 | 1.1 | 95.6 |
| Masuke 2023<sup>116</sup> | 12.0 | - | - | 10.0 | 90.0 | - | 90.0 |
| Mody 2021<sup>117</sup> | 12.0 | - | 40.1 | 40.1 | 59.9 | - | 59.9 |
| Mugenyi 2022<sup>19</sup> | 6.0 | 2.6 | 20.4 | 23.0 | 77.0 | - | 77.0 |
| Mwamuye 2022<sup>118</sup> | 24.0 | 3.8 | 30.3 | 43.6 | 56.4 | 9.5 | 46.9 |
| Nimwesiga 2023<sup>119</sup> | 9.0 | 1.2 | 18.8 | 65.0 | 35.0 | 45 | -10.0 |
| Ojiambo 2023<sup>120</sup> | 9.0 | 9.1 | 55.2 | 64.3 | 35.7 | - | 35.7 |
| Ross 2023<sup>121</sup> | 12.0 | 3.8 | 25.1 | 35.7 | 64.3 | 6.7 | 57.6 |
| Singh 2023<sup>122</sup> | 6.0 | - | 8.1 | 8.1 | 91.9 | - | 91.9 |
| Tlhajoane 2021<sup>34</sup> | 6.0 | 4.5 | 22.7 | 35.5 | 64.5 | 8.3 | 56.2 |
| Zhong 2023<sup>123</sup> | 12.0 | 3.9 | 0.7 | 4.6 | 95.4 | - | 95.4 |
| Simple Averages | 13.4 | 2.9 | 22.1 | 28.1 | 71.9 | 8.1 | 67.6 |
| Weighted Averages<sup>h</sup> | 14.5 | 0.3 | 3.7 | 0.0 | 26.3 | 0.7 | 1.5 |

\*Median Follow-up period by group was reported as: Enrolled in 2016: 24.1 and Enrolled in 2017: 16.6; A collapsed value could not be calculated as Ns by group were not reported
“—” indicates that these data could not be determined from the report.
<sup>a</sup>Calculations: D = 1 - C; F = D - E.
<sup>b</sup>Includes 20 participants who are 10-19 years of age
<sup>c</sup>Mean follow-up time was used
<sup>d</sup>Median follow-up time is based on full cohort
<sup>e</sup>Includes 37 participants who are < 20 years of age
<sup>f</sup>Median follow-up time is based on full cohort which includes the analysis comparing those who are treatment experienced
<sup>g</sup>Includes 11 patients on second-line ART
<sup>h</sup>Weighted by cohort size.

### Quality assessment

The possible range of scores was −7 to 13, with a higher score indicating better quality. The average score for quality was 8.0 (standard deviation (SD): 3.5) with the highest score being 13 and the lowest score being −1, indicating that overall study quality was moderate to high. A majority of studies included a clear and well-defined definition for LTFU. Few studies clearly reported the median follow-up time therefore follow-up time was typically determined based on assumptions described in the methods section above. As a result, this question had the poorest score with a mean of −0.5. It is important to remember that we included abstracts from conferences as well as published studies, and abstracts tended to score on the lower end of the scale due to word limit constraints.

### Publication bias and sensitivity analyses

Figure 4 illustrates retention by last reported timepoint utilizing weighted averages of the interpolated data shows potential publication bias. Studies with shorter follow-up had more attrition than studies with longer follow-up periods. For example, studies reporting to only 12 months had an average retention of 70.2% whereas studies reporting to 24 months had a 12-month average retention of 90.0%. If studies that reported retention only to 12-months had continued follow-up, they likely would have had poorer 24-month retention than studies that followed participants to 24 months. However, we note that this could also be due to varying definitions in loss to follow-up where patients could be allowed to return to care and longer periods of follow-up would increase opportunities for re-engagement.

**Figure 4.**
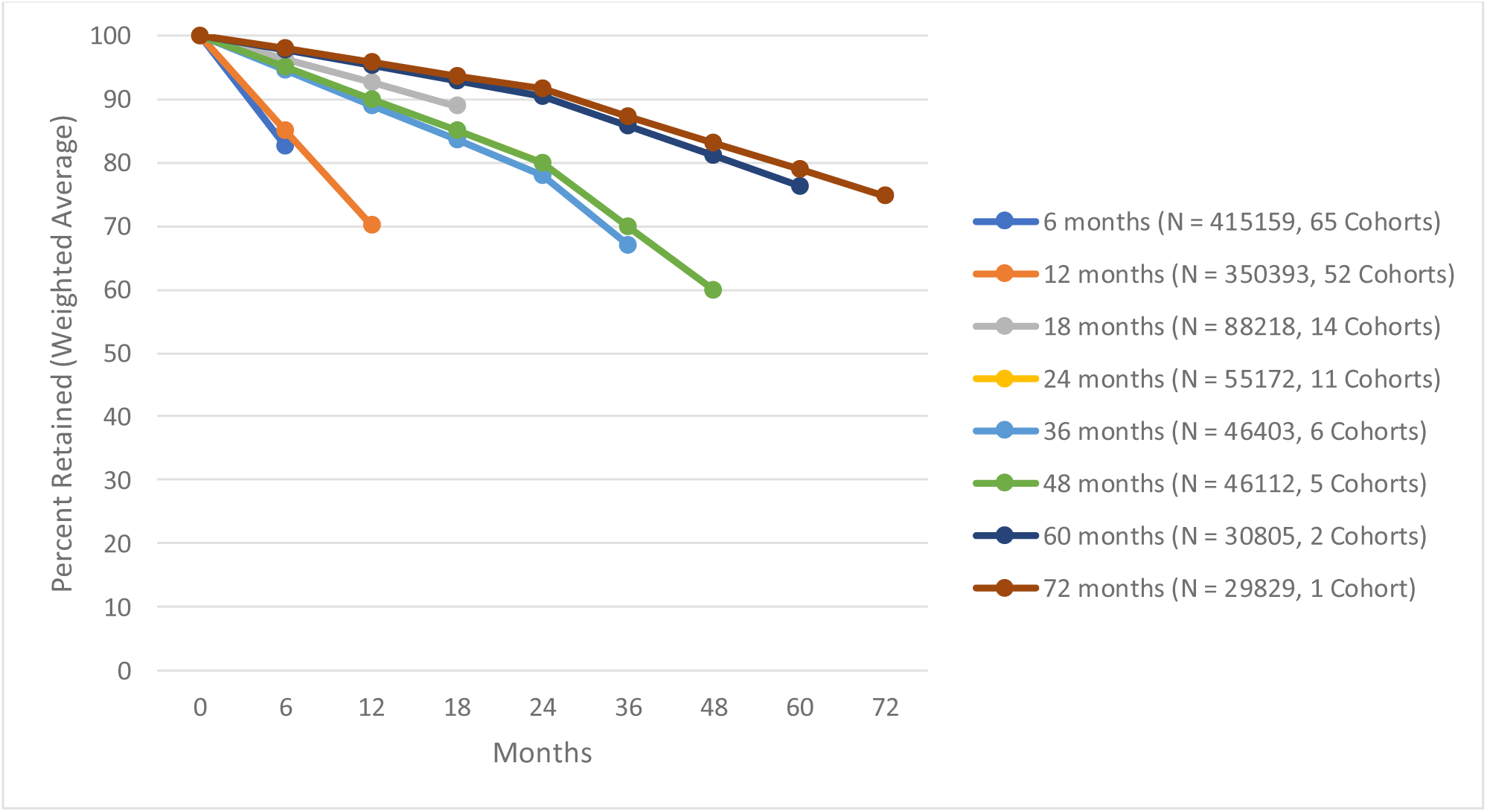
Retention by last reported timepoint utilizing weighted averages of the interpolated data.

Given the possibility of publication bias, we conducted a sensitivity analysis where we modelled attrition under the best-case, worst-case, and midpoint scenarios (Supplemental Figure 2). There is minimal difference in retention rates across the three scenarios in the first year of treatment. The difference widens at 18 months and continues to expand to 92 months. At 24 months, the midpoint scenario retention is 59.5% (best to worst-case range 72.9% to 46.1%). Findings from this analysis differ from our prior reviews where we found minimal difference in retention rates across the three scenarios in the first 24 months.

## DISCUSSION

In this review of retention in the UTT era, capturing 415,159 participants from 28 countries, we estimated pooled retention to be 86.0% at 6 months, 80.0% at 12 months, 82.0% at 24 months, 71.0% at 36 months, and 57.0% at 48 months. These results are largely similar but not identical to findings from our 2015 review, where retention was 91.0% at 6 months, 86.0% at 12 months, 79.0% at 24 months, 75% at 36 months, and 69% at 48 months.

Findings from this updated review highlight a persistent, concerning trend of increasing attrition, especially after the first two years after ART initiation. Our estimated pooled retention rates in the first 12 months following ART initiation in the UTT era were lower compared to the 6- and 12-month retention rates estimated in our prior review. While retention at 24 months was found to be slightly higher in the UTT era compared to in our prior review, retention after 24 months declined drastically and was lower, on average, than in our 2008-2013 estimates. These findings suggest that while UTT resulted in substantial increases in the number of people initiating ART and reduced time to ART initiation, it did not have a large or consistent impact on retention in care after ART initiation. We note that in the pre-UTT era, patients who initiated ART had already demonstrated the willingness and ability to make multiple clinic visits and remain in HIV care. Patients who are initiating in the post-UTT era may thus be different from the patients who returned to the clinic to initiate treatment in the pre-UTT era.^36^

Within our search, very few cohorts reported on retention beyond 36 months. While short-term outcomes are important to evaluate, patients need to be engaged in care and adherent to lifelong ART in order to prevent viral rebound and onward transmission. Notably, retention rates beyond 36 months were concerningly low in our review. Enhanced reporting of long-term retention is essential to improve our understanding of retention barriers across the life course and ultimately identify areas for targeted interventions to improve treatment outcomes. We also note that many cohorts did not report outcomes by year of treatment initiation, which forced us to exclude them from this review; more comprehensive descriptions of published cohorts would improve our understanding of long-term retention. ^37–48^

While our review included all low- and middle-income countries, we found very few studies reporting retention outcomes in countries outside of sub-Saharan Africa. Even within sub-Saharan Africa, the majority of studies came from South Africa. While the majority of PLWH reside on the African continent, it is concerning that we found very few studies reporting on treatment outcomes in other regions. We identified no studies that met our inclusion criteria from Latin America and only one study each from the Caribbean (Haiti) and Europe (Ukraine). There were very few countries in Asia that had retention outcomes in the post-UTT era, and the majority came from Thailand (n=3). Limited reporting on retention from varying geographic regions limits our ability to fully understand the impact of the implementation of UTT within and across regions.

There was considerable variability in how each study defined loss to follow-up and could include undisclosed treatment stoppage, transfer from care, and/or death. Prior studies have demonstrated the definition of loss to follow-up used can meaningfully change retention rates.^49–52^ For example, some cohorts considered LTFU to have occurred at the point their definition of LTFU was met, not allowing participants to return to care, while others allowed participants to return at later time point. Traditional on/off LTFU definitions do not accurately portray the true nature of care engagement as many participants will disengage from care and then re-engage months, if not years, later.^51,53–58^ In addition, participants reported as loss to follow-up in one cohort may have transferred care to another without informing the sending clinic and may present as new participants to the new clinic. This can lead to substantial underestimates of retention.^36,51,59^ Our retention estimates are therefore more representative of “retention at site,” and true rate of retention within ART programs may be much higher.

This review had several limitations. First, larger cohorts (e.g., South Africa, Zambia, and Burundi) likely had a strong impact on our retention estimates, especially when using the interpolated data. Second, while our interpolated data assumed a linear trend between the timepoints, attrition is not always linear especially in the first year of initiation. Any studies where we interpolated the 6- and 12-month outcomes could therefore overestimate retention at the earlier timepoints. Third, although we were interested in understanding retention after UTT adoption, several additional guideline changes (e.g., introduction of dolutegravir for first-line ART and differentiated service delivery models) were also implemented during the study period. As such, observed retention trends reflect a variety of forces and not solely UTT. ^15^ Fourth, while we conducted snowball sampling as well as a hand search to identify any articles that were not MeSH-indexed, some studies reporting on retention were likely missed because the terms utilized to index ART retention are not uniform across publications. The effect of this bias is unknown. Fifth, as few countries outside of the African region were identified, we were unable to evaluate differences across regions. Our findings are likely not indicative of retention outcomes in regions with concentrated epidemics and/or varying treatment guidelines. Sixth, COVID-19 lockdowns and changes in clinic protocols in 2020 may have had an impact on the number of studies being published on retention and on long-term retention outcomes. Finally, our results indicate that there is evidence of publication bias, where cohorts with worse rates of retention are likely to be underrepresented since they did not report on longer-term retention outcomes.

## CONCLUSIONS

In this systematic review, we estimated retention on site in the era of UTT to be 86.0% at 6 months, 80.0% at 12 months, 82.0% at 24 months, and 71.0% at 36 months, and 57.0% at 48 months, suggesting that attrition after the first two years in care remain a concern and continued efforts should be made to ensure participants remain engaged in care over their lifetime. A recent review by Burke et al., based primarily on results from before the COVID-19 pandemic, identified some of the most common reasons for disengagement from treatment in the UTT era, including medication side effects, socioeconomic factors such as employment, and the perception that ART medication lacks benefit.^60^ These findings underscore barriers beyond access to ART and emphasize the need for multilevel interventions to prevent attrition and improve treatment outcomes.^6,61,62^ Furthermore, the COVID-19 pandemic may have impacted retention in care, though the direction of this effect may have varied with some patients more likely to experience treatment interruption due to travel restrictions and clinic closures and others benefiting from pandemic-driven innovations, such as 12-month scripting of ART in South Africa.^63,64^ Future research is needed to assess the impact of the COVID-19 pandemic on retention rates and the extent to which pandemic-induced changes remain today.

## Data Availability

All data produced in the present study are available upon reasonable request to the authors

## References

1. Havlir D, Lockman S, Ayles H, Larmarange J, Chamie G. What do the Universal Test and Treat trials tell us about the path to HIV epidemic control? Collins Iwuji. 2020;10. doi:10.1002/jia2.25455/full

2. Odediran OO, Odukoya OO, Balogun MR, Colasanti JA, Akanmu AS. A Qualitative Study Exploring Factors Associated with Retention in HIV Care among Women with HIV in a Large HIV Clinic in Lagos, Nigeria, after Implementing the Test and Treat Policy. AIDS Res Treat. 2022;2022. doi:10.1155/2022/9074844

3. Yehia BR, Stewart L, Momplaisir F, et al. Barriers and facilitators to patient retention in HIV care. BMC Infect Dis. 2015;15(1). doi:10.1186/s12879-015-0990-0

4. Zinski A, Westfall AO, Gardner LI, et al. The Contribution of Missed Clinic Visits to Disparities in HIV Viral Load Outcomes. Am J Public Health. 2015;105(10):2068–2075. doi:10.2105/AJPH.2015.302695

5. UNAIDS. Global AIDS Strategy 2021-2026.; 2021.

6. Hodgson I, Plummer ML, Konopka SN, et al. A systematic review of individual and contextual factors affecting ART initiation, adherence, and retention for HIV-infected pregnant and postpartum women. PLoS One. 2014;9(11). doi:10.1371/journal.pone.0111421

7. Makurumidze R, Decroo T, Jacobs BKM, et al. Attrition one year after starting antiretroviral therapy before and after the programmatic implementation of HIV “Treat All” in Sub-Saharan Africa: a systematic review and meta-analysis. BMC Infect Dis. 2023;23(1). doi:10.1186/s12879-023-08551-y

8. Fox MP, Rosen S. Retention of Adult Patients on Antiretroviral Therapy in Low- and Middle-Income Countries: Systematic Review and Meta-analysis 2008-2013. J Acquir Immune Defic Syndr. 2015;69(1):98–108. doi:10.1097/QAI.0000000000000553

9. Rosen S, Fox MP, Gill CJ. Patient retention in antiretroviral therapy programs in sub-Saharan Africa: a systematic review. PLoS Med. 2007;4(10):e298. doi:10.1371/journal.pmed.0040298

10. Long L, Kuchukhidze S, Pascoe S, et al. Retention in care and viral suppression in differentiated service delivery models for HIV treatment delivery in sub-Saharan Africa: a rapid systematic review. J Int AIDS Soc. 2020;23(11). doi:10.1002/jia2.25640

11. Hagey JM, Li X, Barr-Walker J, et al. Differentiated HIV care in sub-Saharan Africa: a scoping review to inform antiretroviral therapy provision for stable HIV-infected individuals in Kenya. AIDS Care. 2018;30(12):1477–1487. doi:10.1080/09540121.2018.1500995

12. Limbada M, Zijlstra G, Macleod D, Ayles H, Fidler S. A systematic review of the effectiveness of non-health facility based care delivery of antiretroviral therapy for people living with HIV in sub-Saharan Africa measured by viral suppression, mortality and retention on ART. BMC Public Health. 2021;21(1):1110. doi:10.1186/s12889-021-11053-8

13. World Health Organization. Consolidated Guidelines on the Use of Antiretroviral Drugs for Treating and Preventing HIV Infection. 2nd Edition.; 2016.

14. World Health Organization. Antiretroviral Therapy for HIV Infection in Adults and Adolescents. Recommendations for a Public Health Approach. 2010 Revision.; 2010.

15. World Health Organization, Hampton G. UPDATED RECOMMENDATIONS ON FIRST-LINE AND SECOND-LINE ANTIRETROVIRAL REGIMENS AND POST-EXPOSURE PROPHYLAXIS AND RECOMMENDATIONS ON EARLY INFANT DIAGNOSIS OF HIV POLICY BRIEF HIV TREATMENT-INTERIM GUIDANCE.; 2018. http://apps.who.int/bookorders.

16. World Health Organization. THE USE OF ANTIRETROVIRAL DRUGS FOR TREATING AND PREVENTING HIV INFECTION 2016.; 2016.

17. Brennan AT, Bor J, Davies MA, et al. Medication side effects and retention in HIV treatment: A regression discontinuity study of tenofovir implementation in South Africa and Zambia. Am J Epidemiol. 2018;187(9):1990–2001. doi:10.1093/aje/kwy093

18. Girum T, Yasin F, Wasie A, Shumbej T, Bekele F, Zeleke B. The effect of “universal test and treat” program on HIV treatment outcomes and patient survival among a cohort of adults taking antiretroviral treatment (ART) in low income settings of Gurage zone, South Ethiopia. AIDS Res Ther. 2020;17(1). doi:10.1186/s12981-020-00274-3

19. Mugenyi L, Nanfuka M, Byawaka J, et al. Effect of universal test and treat on retention and mortality among people living with HIV-infection in Uganda: An interrupted time series analysis. PLoS One. 2022;17(5 May). doi:10.1371/journal.pone.0268226

20. Bekolo CE, Ndeso SA, Gougue CP, et al. The effect of the Universal Test and Treat policy uptake on CD4 count testing and incidence of opportunistic infections among people living with HIV infection in Cameroon: a retrospective analysis of routine data. Dialogues in Health. 2023;2. doi:10.1016/j.dialog.2023.100120

21. Onoya D, Hendrickson C, Sineke T, et al. Attrition in HIV care following HIV diagnosis: a comparison of the pre-UTT and UTT eras in South Africa. J Int AIDS Soc. 2021;24(2). doi:10.1002/jia2.25652

22. Nawfal ES, Gray A, Sheehan DM, Ibañez GE, Trepka MJ. A Systematic Review of the Impact of HIV-Related Stigma and Serostatus Disclosure on Retention in Care and Antiretroviral Therapy Adherence Among Women with HIV in the United States/Canada. AIDS Patient Care STDS. 2024;38(1):23–49. doi:10.1089/apc.2023.0178

23. Birhanu MY, Bekele GM, Yirdaw G, Demissie BS, Getahun GK, Jemberie SS. Incidence and predictors of loss to follow-up among Ethiopian children on antiretroviral therapy: a systematic review and meta-analysis. BMC Public Health. 2024;24(1):328. doi:10.1186/s12889-024-17778-6

24. The World Bank. The World by Income and Region. 2023. 2023. Accessed August 15, 2024. https://datatopics.worldbank.org/world-development-indicators/the-world-by-income-and-region.html

25. Benade M, Maskew M, Juntunen A, Flynn DB, Rosen S. Prior exposure to antiretroviral therapy among adult patients presenting for HIV treatment initiation or reinitiation in sub-Saharan Africa: a systematic review. BMJ Open. 2023;13(11):e071283. doi:10.1136/bmjopen-2022-071283

26. Sayers A. Tips and tricks in performing a systematic review. Br J Gen Pract. 2007;57(538):425. http://www.ncbi.nlm.nih.gov/pubmed/17504612

27. COVIDENCE. Covidence - Better systematic review management. Accessed March 18, 2024. https://www.covidence.org/

28. Munn Z, Moola S, Lisy K, Riitano D, Tufanaru C. Chapter 5: Systematic Reviews of Prevalence and Incidence. In: Aromatris E, Munn Z, eds. JBI Manual for Evidence Synthesis. JBI; 2020.

29. Wells G, Shea B, O’Connell D, et al. The Newcastle-Ottawa Scale (NOS) for assessing the quality of nonrandomized studies in meta-analyses. Department of Epidemiology and Community Medicine, University of Ottawa. Accessed March 18, 2024. https://www.ohri.ca/programs/clinical_epidemiology/oxford.asp

30. Byamukama A, Golding PM. Predictors of mortality among people living with HIV in the test and treat era within rural Uganda: a retrospective cohort study. African Journal of AIDS Research. 2022;21(3):231–238. doi:10.2989/16085906.2022.2056062

31. Dorward J, Sookrajh Y, Gate K, et al. HIV treatment outcomes among people with initiation CD4 counts >500 cells/µL after implementation of Treat All in South African public clinics: a retrospective cohort study. J Int AIDS Soc. 2020;23(4). doi:10.1002/jia2.25479

32. Lin L, Xu C. Arcsine-based transformations for meta-analysis of proportions: Pros, cons, and alternatives. Health Sci Rep. 2020;3(3). doi:10.1002/hsr2.178

33. Lin L, Chu H. Meta-analysis of proportions using generalized linear mixed models. Epidemiology. 2020;31(5):713–717. doi:10.1097/EDE.0000000000001232

34. Tlhajoane M, Dzamatira F, Kadzura N, Nyamukapa C, Eaton JW, Gregson S. Incidence and predictors of attrition among patients receiving ART in eastern Zimbabwe before, and after the introduction of universal ‘treat-all’ policies: A competing risk analysis. PLOS Global Public Health. 2021;1(10). doi:10.1371/journal.pgph.0000006

35. Ibiloye O, Decroo T, Eyona N, Eze P, Agada P. Characteristics and early clinical outcomes of key populations attending comprehensive community-based HIV care: Experiences from Nasarawa State, Nigeria. PLoS One. 2018;13(12). doi:10.1371/journal.pone.0209477

36. Fox MP, Rosen S. A new cascade of HIV care for the era of “treat all.” PLoS Med. 2017;14(4). doi:10.1371/journal.pmed.1002268

37. Linn KZ, Shewade HD, Htet KKK, Maung TM, Hone S, Oo HN. Time to anti-retroviral therapy among people living with HIV enrolled into care in Myanmar: how prepared are we for ‘test and treat’? Glob Health Action. 2018;11(1):1520473. doi:10.1080/16549716.2018.1520473

38. Aung ZZ, Oo MM, Tripathy JP, et al. Are death and loss to follow-up still high in people living with HIV on ART after national scale-up and earlier treatment initiation? A large cohort study in government hospitalbased setting, Myanmar: 2013-2016. PLoS One. 2018;13(9). doi:10.1371/journal.pone.0204550

39. Aung ZZ, Oo MM, Tripathy JP, et al. Are death and loss to follow-up still high in people living with HIV on ART after national scale-up and earlier treatment initiation? A large cohort study in government hospitalbased setting, Myanmar: 2013-2016. PLoS One. 2018;13(9). doi:10.1371/journal.pone.0204550

40. Carriquiry G, Giganti MJ, Castilho JL, et al. Virologic failure and mortality in older ART initiators in a multisite Latin American and Caribbean Cohort. J Int AIDS Soc. 2018;21(3). doi:10.1002/jia2.25088

41. Teasdale CA, Yuengling K, Preko P, et al. Persons living with advanced HIV disease: need for novel care models. J Int AIDS Soc. 2018;21(12). doi:10.1002/jia2.25210

42. Nguyen HH, Bui DD, Dinh TT, et al. A prospective “test-and-treat” demonstration project among people who inject drugs in Vietnam. J Int AIDS Soc. 2018;21(7). doi:10.1002/jia2.25151

43. Huang YC, Sun HY, Chuang YC, et al. Short-term outcomes of rapid initiation of antiretroviral therapy among HIV-positive patients: Real-world experience from a single-centre retrospective cohort in Taiwan. BMJ Open. 2019;9(9). doi:10.1136/bmjopen-2019-033246

44. Aemro A, Wassie M, Chekol B. Incidence and predictors of mortality within the first year of antiretroviral therapy initiation at Debre-Markos Referral Hospital, Northwest Ethiopia: A retrospective follow up study. PLoS One. 2021;16(5 May). doi:10.1371/journal.pone.0251648

45. Limmade Y, Fransisca L, Rodriguez-Fernandez R, Bangs MJ, Rothe C. HIV treatment outcomes following antiretroviral therapy initiation and monitoring: A workplace program in Papua, Indonesia. PLoS One. 2019;14(2). doi:10.1371/journal.pone.0212432

46. Ayeno HD, Imer AA. First-line antiretroviral treatment failure and associated factors in HIV patients following highly active antiretroviral therapy at the Shashemene Referral Hospital, Oromia region, Ethiopia. HIV and AIDS Review. 2020;19(2):125–131. doi:10.5114/hivar.2020.96388

47. Wu G, Zhou C, Zhang X, et al. Higher risks of virologic failure and all-cause deaths among older people living with hiv in chongqing, China. AIDS Res Hum Retroviruses. 2019;35(11-12):1095–1102. doi:10.1089/aid.2019.0096

48. Mwe Nom NA, Kyaw KWY, Kumar AMV, et al. HIV care cascade among prisoners of the Mandalay Central Prison in Myanmar: 2011–2018. Trop Med Infect Dis. 2020;5(1). doi:10.3390/tropicalmed5010004

49. van der Kop ML, Nagide PI, Thabane L, et al. Retention in clinic versus retention in care during the first year of HIV care in Nairobi, Kenya: a prospective cohort study. J Int AIDS Soc. 2018;21(11). doi:10.1002/jia2.25196

50. Suffrin JCD, Rosenthal A, Kamtsendero L, et al. Re-engagement and retention in HIV care after preventive default tracking in a cohort of HIV-infected patients in rural Malawi: A mixed-methods study. Togun TO, ed. PLOS Global Public Health. 2024;4(2):e0002437. doi:10.1371/journal.pgph.0002437

51. Fox MP, Bor J, Brennan AT, et al. Estimating retention in HIV care accounting for patient transfers: A national laboratory cohort study in South Africa. PLoS Med. 2018;15(6). doi:10.1371/journal.pmed.1002589

52. Grimsrud AT, Cornell M, Egger M, Boulle A, Myer L. Impact of definitions of loss to follow-up (LTFU) in antiretroviral therapy program evaluation: Variation in the definition can have an appreciable impact on estimated proportions of LTFU. J Clin Epidemiol. 2013;66(9):1006–1013. doi:10.1016/j.jclinepi.2013.03.013

53. Keene CM, Ragunathan A, Euvrard J, et al. Measuring patient engagement with HIV care in sub-Saharan Africa: a scoping study. J Int AIDS Soc. 2022;25(10):e26025. doi:10.1002/jia2.26025

54. Ehrenkranz P, Rosen S, Boulle A, et al. The revolving door of HIV care: Revising the service delivery cascade to achieve the UNAIDS 95-95-95 goals. PLoS Med. 2021;18(5):e1003651. doi:10.1371/journal.pmed.1003651

55. Etoori D, Kabudula CW, Wringe A, et al. Investigating clinic transfers among HIV patients considered lost to follow-up to improve understanding of the HIV care cascade: Findings from a cohort study in rural north-eastern South Africa. PLOS Global Public Health. 2022;2(5):e0000296. doi:10.1371/journal.pgph.0000296

56. Bengtson AM, Espinosa Dice AL, Kirwa K, Cornell M, Colvin CJ, Lurie MN. Patient Transfers and Their Impact on Gaps in Clinical Care: Differences by Gender in a Large Cohort of Adults Living with HIV on Antiretroviral Therapy in South Africa. AIDS Behav. 2021;25(10):3337–3346. doi:10.1007/s10461-021-03191-2

57. Espinosa Dice AL, Bengtson AM, Mwenda KM, Colvin CJ, Lurie MN. Quantifying clinic transfers among people living with HIV in the Western Cape, South Africa: a retrospective spatial analysis. BMJ Open. 2021;11(12):e055712. doi:10.1136/bmjopen-2021-055712

58. Pry JM, Mwila C, Kapesa H, et al. Estimating potential silent transfer using baseline viral load measures among people presenting as new to HIV care in Lusaka, Zambia: a cross-sectional study. BMJ Open. 2023;13(5):e070384. doi:10.1136/bmjopen-2022-070384

59. Edwards JK, Lesko CR, Herce ME, et al. Gone But Not Lost: Implications for Estimating HIV Care Outcomes When Loss to Clinic Is Not Loss to Care. Epidemiology. 2020;31(4):570–577. doi:10.1097/EDE.0000000000001201

60. Burke RM, Rickman HM, Pinto C, Ehrenkranz P, Choko A, Ford N. Reasons for disengagement from antiretroviral care in the era of “Treat All” in low- or middle-income countries: a systematic review. J Int AIDS Soc. 2024;27(3). doi:10.1002/jia2.26230

61. Casale M, Carlqvist A, Cluver L. Recent Interventions to Improve Retention in HIV Care and Adherence to Antiretroviral Treatment Among Adolescents and Youth: A Systematic Review. AIDS Patient Care STDS. 2019;33(6):237–252. doi:10.1089/apc.2018.0320

62. Govindasamy D, Meghij J, Kebede Negussi E, Clare Baggaley R, Ford N, Kranzer K. Interventions to improve or facilitate linkage to or retention in pre-ART (HIV) care and initiation of ART in low- and middle-income settings--a systematic review. J Int AIDS Soc. 2014;17(1):19032. doi:10.7448/IAS.17.1.19032

63. Lewis L, Sookrajh Y, van der Molen J, et al. Clinical outcomes after extended 12-month antiretroviral therapy prescriptions in a community-based differentiated HIV service delivery programme in South Africa: a retrospective cohort study. J Int AIDS Soc. 2023;26(9). doi:10.1002/jia2.26164

64. Deghaye N, Nkonki L, Rensburg R, Van Schalkwyk C. Examining the Unintended Consequences of the COVID-19 Pandemic on Public Sector Health Facility Visits: The First 150 Days.; 2020.

65. Ahmed I, Demissie M, Worku A, Gugsa S, Berhane Y. Effectiveness of same-day antiretroviral therapy initiation in retention outcomes among people living with human immunodeficiency virus in Ethiopia: empirical evidence. BMC Public Health. 2020;20(1). doi:10.1186/s12889-020-09887-9

66. Alhaj M, Amberbir A, Singogo E, et al. Retention on antiretroviral therapy during Universal Test and Treat implementation in Zomba district, Malawi: a retrospective cohort study. J Int AIDS Soc. 2019;22(2):e25239. doi:10.1002/jia2.25239

67. Atuhaire L, Shumba CS, Mapahla L, Nyasulu PS. A retrospective cross sectional study assessing factors associated with retention and non-viral suppression among HIV positive FSWs receiving antiretroviral therapy from primary health care facilities in Kampala, Uganda. BMC Infect Dis. 2022;22(1). doi:10.1186/s12879-022-07614-w

68. Avalos A, Gaolathe T, Brown D, et al. 12 Month Outcomes on Dolutegravir Based Regimens in Botswana: The BEAT Cohort Study. Conference on Retroviruses and Opportunistic Infections. Published online March 2019.

69. Benzekri NA, Sambou JF, Ndong S, et al. Food insecurity predicts loss to follow-up among people living with HIV in Senegal, West Africa. AIDS Care - Psychological and Socio-Medical Aspects of AIDS/HIV. 2022;34(7):878–886. doi:10.1080/09540121.2021.1894316

70. Cassidy T, Cornell M, Makeleni B, et al. Attrition from Care Among Men Initiating ART in Male-Only Clinics Compared with Men in General Primary Healthcare Clinics in Khayelitsha, South Africa: A Matched Propensity Score Analysis. AIDS Behav. 2023;27(1):358–369. doi:10.1007/s10461-022-03772-9

71. Chauke P, Huma M, Madiba S. Lost to follow up rate in the first year of art in adults initiated in a universal test and treat programme: A retrospective cohort study in ekurhuleni district, south africa. Pan African Medical Journal. 2020;37:1–13. doi:10.11604/pamj.2020.37.198.25294

72. Dumchev K, Kiriazova T, Riabokon S, et al. Comparative Clinical Outcomes With Scale-up of Dolutegravir as First-Line Antiretroviral Therapy in Ukraine. J Acquir Immune Defic Syndr. 2022;91(2):197–209. doi:10.1097/QAI.0000000000003038

73. Eamsakulrat P, Kiertiburanakul S. The Impact of Timing of Antiretroviral Therapy Initiation on Retention in Care, Viral Load Suppression and Mortality in People Living with HIV: A Study in a University Hospital in Thailand. J Int Assoc Provid AIDS Care. 2022;21. doi:10.1177/23259582221082607

74. Ntamatungiro AJ, Eichenberger A, Okuma J, et al. Transitioning to Dolutegravir in a Programmatic Setting: Virological Outcomes and Associated Factors Among Treatment-Naive Patients With HIV-1 in the Kilombero and Ulanga Antiretroviral Cohort in Rural Tanzania. Open Forum Infect Dis. 2023;10(7). doi:10.1093/ofid/ofad321

75. Eshiwani SJ, Mutal K, Muiruri P, Kubo E. A comparison of outcomes of same day versus delayed Antiretroviral treatment (ART) initiation in the “Test and Treat era” in Kenyatta National Hospital. International AIDS Conference. Published online 2018.

76. Harooni MZ, Atarud AA, Ehsan E, Alokozai A, McFarland W, Mirzazadeh A. Gaps in the continuum of care among people living with HIV in Afghanistan. Int J STD AIDS. 2022;33(3):282–288. doi:10.1177/09564624211055299

77. Hirasen K, Fox MP, Hendrickson CJ, Sineke T, Onoya D. HIV treatment outcomes among patients initiated on antiretroviral therapy pre and post-universal test and treat guidelines in South Africa. Ther Clin Risk Manag. 2020;16:169–180. doi:10.2147/TCRM.S227290

78. Hoang NT, Foo TJ, Tran BX, et al. Structural barriers for retention of HIV/AIDS patients after initiating antiretroviral therapy in outpatient clinics of Vietnam. AIDS Care - Psychological and Socio-Medical Aspects of AIDS/HIV. 2022;34(8):992–999. doi:10.1080/09540121.2021.1929816

79. Ibiloye O, Jwanle P, Masquillier C, et al. Long-term retention and predictors of attrition for key populations receiving antiretroviral treatment through communitybased ART in Benue State Nigeria: A retrospective cohort study. PLoS One. 2021;16(11). doi:10.1371/journal.pone.0260557

80. Izudi J, Kiragga AN, Kalyesubula P, Okoboi S, Castelnuovo B. Effect of the COVID-19 pandemic restrictions on outcomes of HIV care among adults in Uganda. Medicine (United States). 2022;101(36):E30282. doi:10.1097/MD.0000000000030282

81. Jamieson L, Rosen S, Phiri B, et al. How soon should patients be eligible for differentiated service delivery models for antiretroviral treatment? Evidence from a retrospective cohort study in Zambia. BMJ Open. 2022;12(12). doi:10.1136/bmjopen-2022-064070

82. Januraga PP, Reekie J, Mulyani T, et al. The cascade of HIV care among key populations in Indonesia: a prospective cohort study. Lancet HIV. 2018;5(10):e560–e568. doi:10.1016/S2352-3018(18)30148-6

83. Johansson M, Penno C, Winqvist N, Tesfaye F, Björkman P. How does HIV testing modality impact the cascade of care among persons diagnosed with HIV in Ethiopia? Glob Health Action. 2021;14(1). doi:10.1080/16549716.2021.1933788

84. Kimanga DO, Oramisi VA, Hassan AS, et al. Uptake and effect of universal test-and-treat on twelve months retention and initial virologic suppression in routine HIV program in Kenya. PLoS One. 2022;17(11 November). doi:10.1371/journal.pone.0277675

85. Lilian RR, Rees K, McIntyre JA, Struthers HE, Peters RPH. Same-day antiretroviral therapy initiation for HIV-infected adults in South Africa: Analysis of routine data. PLoS One. 2020;15(1). doi:10.1371/journal.pone.0227572

86. Makurumidze R, Buyze J, Decroo T, et al. Patient-mix, programmatic characteristics, retention and predictors of attrition among patients starting antiretroviral therapy (ART) before and after the implementation of HIV “treat All” in Zimbabwe. PLoS One. 2020;15(10 October). doi:10.1371/journal.pone.0240865

87. Matare T, Shewade HD, Ncube RT, et al. Anti-retroviral therapy after “Treat All” in Harare, Zimbabwe: What are the changes in uptake, time to initiation and retention? F1000Res. 2020;9:287. doi:10.12688/f1000research.23417.1

88. Matthews LT, Orrell C, Bwana MB, et al. Adherence to HIV antiretroviral therapy among pregnant and postpartum women during the Option B+ era: 12-month cohort study in urban South Africa and rural Uganda. J Int AIDS Soc. 2020;23(8). doi:10.1002/jia2.25586

89. Mayasi N, Situakibanza H, Mbula M, et al. Retention in care and predictors of attrition among HIV-infected patients who started antiretroviral therapy in Kinshasa, DRC, before and after the implementation of the ‘treat-all’ strategy. PLOS Global Public Health. 2022;2(3):e0000259. doi:10.1371/journal.pgph.0000259

90. Mshweshwe-Pakela N, Hansoti B, Mabuto T, et al. Feasibility of implementing same-day antiretroviral therapy initiation during routine care in Ekurhuleni District, South Africa: Retention and viral load suppression. South Afr J HIV Med. 2020;21(1). doi:10.4102/SAJHIVMED.V21I1.1085

91. Nshimirimana C, Ndayizeye A, Smekens T, Vuylsteke B. Loss to follow-up of patients in HIV care in Burundi: A retrospective cohort study. Tropical Medicine and International Health. 2022;27(6):574–582. doi:10.1111/tmi.13753

92. Onoya D, Sineke T, Hendrickson C, et al. Impact of the test and treat policy on delays in antiretroviral therapy initiation among adult HIV positive patients from six clinics in Johannesburg, South Africa: Results from a prospective cohort study. BMJ Open. 2020;10(3). doi:10.1136/bmjopen-2019-030228

93. Opio D, Semitala FC, Kakeeto A, et al. Loss to follow-up and associated factors among adult people living with HIV at public health facilities in Wakiso district, Uganda: A retrospective cohort study. BMC Health Serv Res. 2019;19(1). doi:10.1186/s12913-019-4474-6

94. Opito R, Mpagi J, Bwayo D, Okello F, Mugisha K, Napyo A. Treatment outcome of the implementation of HIV test and treat policy at the AIDs Support Organization (TASO) Tororo clinic, Eastern Uganda: A retrospective cohort study. PLoS One. 2020;15(9 September). doi:10.1371/journal.pone.0239087

95. Pillay P, Wadley AL, Cherry CL, Karstaedt AS, Kamerman PR. Clinical diagnosis of sensory neuropathy in HIV patients treated with tenofovir: A 6-month follow-up study. Journal of the Peripheral Nervous System. 2019;24(4):304–313. doi:10.1111/jns.12349

96. Ajeh RA, Gregory HE, Thomas EO, et al. Determinants of retention in hiv antiretroviral treatment (Art) in the cameroon international epidemiology database to evaluate aids (iedea) study clinics: The context of the hiv treat all strategy in cameroon. Pan African Medical Journal. 2021;40. doi:10.11604/pamj.2021.40.129.22642

97. Romo ML, Brazier E, Mahambou-Nsondé D, et al. Real-world use and outcomes of dolutegravir-containing antiretroviral therapy in HIV and tuberculosis co-infection: a site survey and cohort study in sub-Saharan Africa. J Int AIDS Soc. 2022;2022:25961. doi:10.1002/jia2.25961/full

98. Ross J, Sinayobye J d’Amour, Yotebieng M, et al. Early outcomes after implementation of treat all in Rwanda: an interrupted time series study. J Int AIDS Soc. 2019;22(4). doi:10.1002/jia2.25279

99. Seekaew P, Janamnuaysook R, Teeratakulpisarn N, et al. Transgender-Led Same-Day Antiretroviral Therapy Services at the Tangerine Community Health Center in Bangkok, Thailand. International AIDS Society Conference. Published online 2019.

100. Seekaew P, Phanuphak N, Teeratakulpisarn N, et al. Same-day antiretroviral therapy initiation hub model at the Thai Red Cross Anonymous Clinic in Bangkok, Thailand: an observational cohort study. J Int AIDS Soc. 2021;24:25869. doi:10.1002/jia2.25869/full

101. Ssempijja V, Namulema E, Ankunda R, et al. Temporal trends of early mortality and its risk factors in HIV-infected adults initiating antiretroviral therapy in Uganda. EClinicalMedicine. 2020;28. doi:10.1016/j.eclinm.2020.100600

102. Stafford KA, Odafe SF, Lo J, et al. Evaluation of the clinical outcomes of the Test and Treat strategy to implement Treat All in Nigeria: Results from the Nigeria multi-center ARt study. PLoS One. 2019;14(7). doi:10.1371/journal.pone.0218555

103. Stockton MA, Gaynes BN, Hosseinipour MC, et al. Association Between Depression and HIV Care Engagement Outcomes Among Patients Newly Initiating ART in Lilongwe, Malawi. AIDS Behav. 2021;25(3):826–835. doi:10.1007/s10461-020-03041-7

104. Teshale AB, Tsegaye AT, Wolde HF. Incidence and predictors of loss to follow up among adult HIV patients on antiretroviral therapy in University of Gondar Comprehensive Specialized Hospital: A competing risk regression modeling. PLoS One. 2020;15(1). doi:10.1371/journal.pone.0227473

105. Bernard Marc J, Pierre S, Rivera V, et al. A Model of Successful ART Initiation in the Context of Massive Civil Unrest in Haiti. Conference on Retroviruses and Opportunistic Infections. Published online 2023. A MODEL OF SUCCESSFUL ART INITIATION IN THE CONTEXT OF MASSIVE CIVIL UNREST IN HAITI

106. Chagomerana MB, Harrington BJ, DiPrete BL, et al. Three-year outcomes for women newly initiated on lifelong antiretroviral therapy during pregnancy – Malawi option B+. AIDS Res Ther. 2023;20(1). doi:10.1186/s12981-023-00523-1

107. Dorward J, Sookrajh Y, Khubone T, et al. Implementation and outcomes of dolutegravir-based first-line antiretroviral therapy for people with HIV in South Africa: a retrospective cohort study. Lancet HIV. 2023;10(5):e284–e294. doi:10.1016/S2352-3018(23)00047-4

108. Gemechu A, Mihret A, Aseffa A, Howe R, Seyoum B, Mulu A. Loss to Follow-up and Death Among Individuals With Newly Diagnosed Human Immunodeficiency Virus Receiving Dolutegravir-Based First-Line Antiretroviral Treatment in Eastern Ethiopia: Implications for 95% United Nations Targets. Open Forum Infect Dis. 2023;10(11). doi:10.1093/ofid/ofad522

109. Govere SM, Kalinda C, Chimbari MJ. The impact of same-day antiretroviral therapy initiation on retention in care and clinical outcomes at four eThekwini clinics, KwaZulu-Natal, South Africa. BMC Health Serv Res. 2023;23(1). doi:10.1186/s12913-023-09801-0

110. Hamooya BM, Mutembo S, Muyunda B, et al. HIV test-and-treat policy improves clinical outcomes in Zambian adults from Southern Province: a multicenter retrospective cohort study. Front Public Health. 2023;11. doi:10.3389/fpubh.2023.1244125

111. Joaquim L, Miranda MNS, Pimentel V, et al. Retention in Care and Virological Failure among Adult HIV-Positive Patients on First-Line Antiretroviral Treatment in Maputo, Mozambique. Viruses. 2023;15(10). doi:10.3390/v15101978

112. Kimanga DO, Makory VNB, Hassan AS, et al. Impact of the COVID-19 pandemic on routine HIV care and antiretroviral treatment outcomes in Kenya: A nationally representative analysis. PLoS One. 2023;18(11 November). doi:10.1371/journal.pone.0291479

113. Lavoie MCC, Ehoche A, Blanco N, et al. Effect of Test and Treat on clinical outcomes in Nigeria: A national retrospective study. PLoS One. 2023;18(8 August). doi:10.1371/journal.pone.0284847

114. MacKellar D, Hlophe T, Ujamaa D, et al. Antiretroviral therapy initiation and retention among clients who received peer-delivered linkage case management and standard linkage services, Eswatini, 2016–2020: retrospective comparative cohort study. Archives of Public Health. 2022;80(1). doi:10.1186/s13690-022-00810-9

115. Masaba RO, Herrera N, Siamba S, et al. Advanced HIV disease in Homa Bay County, Kenya: Characteristics of newly-diagnosed and antiretroviral therapy-experienced clients. Medicine (United States). 2023;102(51). doi:10.1097/MD.0000000000036716

116. Masuke R, Kihaga Y, Mashala M, Ndalio S, Sukari O, Panga O. Twelve months antiretroviral therapy retention among clients newly enrolled to care and treatment services in Geita region, Tanzania: does universal test and treat matter? Pan African Medical Journal. 2023;46. doi:10.11604/pamj.2023.46.20.40772

117. Mody A, Sikazwe I, Namwase AS, et al. Effects of implementing universal and rapid HIV treatment on initiation of antiretroviral therapy and retention in care in Zambia: a natural experiment using regression discontinuity. Lancet HIV. 2021;8(12):e755–e765. doi:10.1016/S2352-3018(21)00186-7

118. Mwamuye IC, Karanja S, Msanzu JB, Adem A, Kerich M, Ngari M. Factors associated with poor outcomes among people living with HIV started on antiretroviral therapy before and after implementation of “test and treat” program in Coastal Kenya. PLoS One. 2022;17(9 September). doi:10.1371/journal.pone.0270653

119. Nimwesiga C, Taremwa IM, Nakanjako D, Nasuuna E. Factors Associated with Retention in HIV Care Among HIV-Positive Adolescents in Public Antiretroviral Therapy Clinics in Ibanda District, Rural South Western Uganda. HIV/AIDS - Research and Palliative Care. 2023;15:71–81. doi:10.2147/HIV.S401611

120. Ojiambo KO, Nakku J, Wangi RN, et al. Socio-demographic and clinical characteristics associated with retenti in care among adults living with HIV and severe mental illness and reasons for loss to follow-up in Uganda: a mixed-methods study. BMJ Open. 2023;13(10). doi:10.1136/bmjopen-2023-073623

121. Ross J, Brazier E, Fatti G, et al. Same-Day Antiretroviral Therapy Initiation as a Predictor of Loss to Follow-up and Viral Suppression Among People With Human Immunodeficiency Virus in Sub-Saharan Africa. Clinical Infectious Diseases. 2023;76(1):39–47. doi:10.1093/cid/ciac759

122. Singh B, Guliani A, Hanumanthu V, et al. A prospective study to estimate the incidence and pattern of adverse drug reactions to first-line antiretroviral therapy (tenofovir, efavirenz, and lamivudine). Indian J Sex Transm Dis AIDS. 2023;44(1):6–10. doi:10.4103/ijstd.ijstd_44_21

123. Zhong M, Li M, Qi M, et al. A retrospective clinical study of dolutegravir-versus efavirenz-based regimen in treatment-naïve patients with advanced HIV infection in Nanjing, China. Front Immunol. 2023;13. doi:10.3389/fimmu.2022.1033098

