## Supplemental Tables and Figures for "Systematic Review and Meta-Analysis of Retention and Disengagement After Initiation on Antiretroviral Therapy in Low- and Middle-Income Countries After the Introduction of Universal Test and Treat Policies"

**Supplemental Figure 1.** Forest plots of retention rates and their corresponding 95% CIs by time point reported to at 18-, 48-, and 60-months

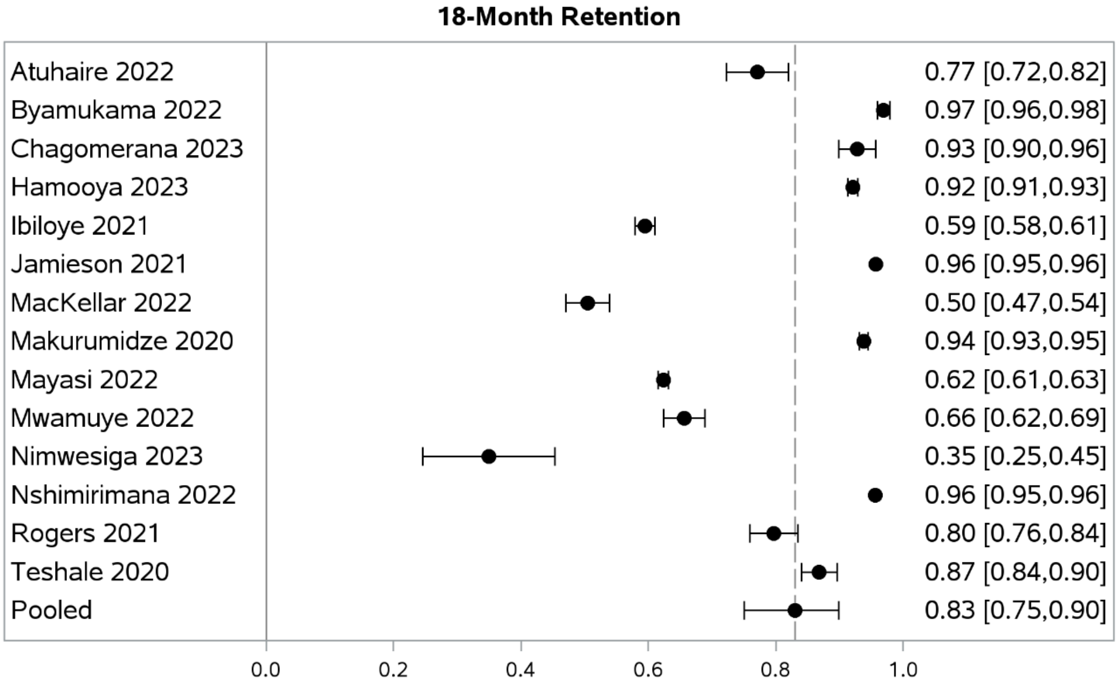


*Dashed line represents the pooled point estimate*
*Interpolated data is used for studies that did not report at that timepoint but did at the timepoint prior and after it*
**Ex. A study reported retention at 12 and 24 months therefore 18-month retention is interpolated*

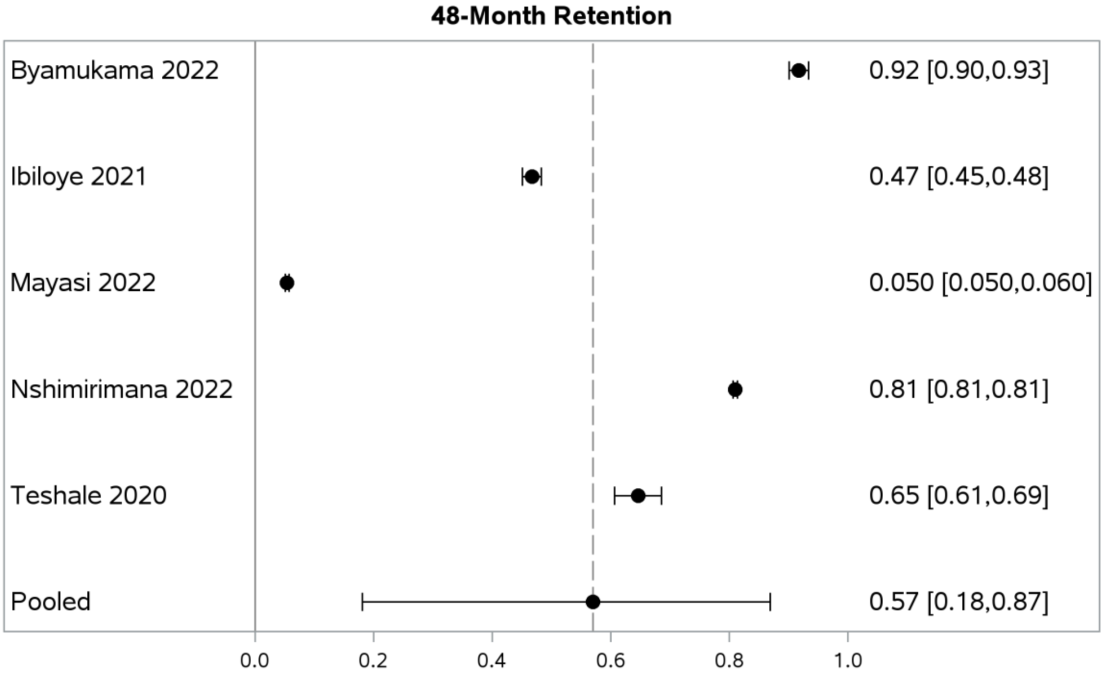


*Dashed line represents the pooled point estimate*
*Interpolated data is used for studies that did not report at that timepoint but did at the timepoint prior and after it*
**Ex. A study reported retention at 12 and 24 months therefore 18-month retention is interpolated*

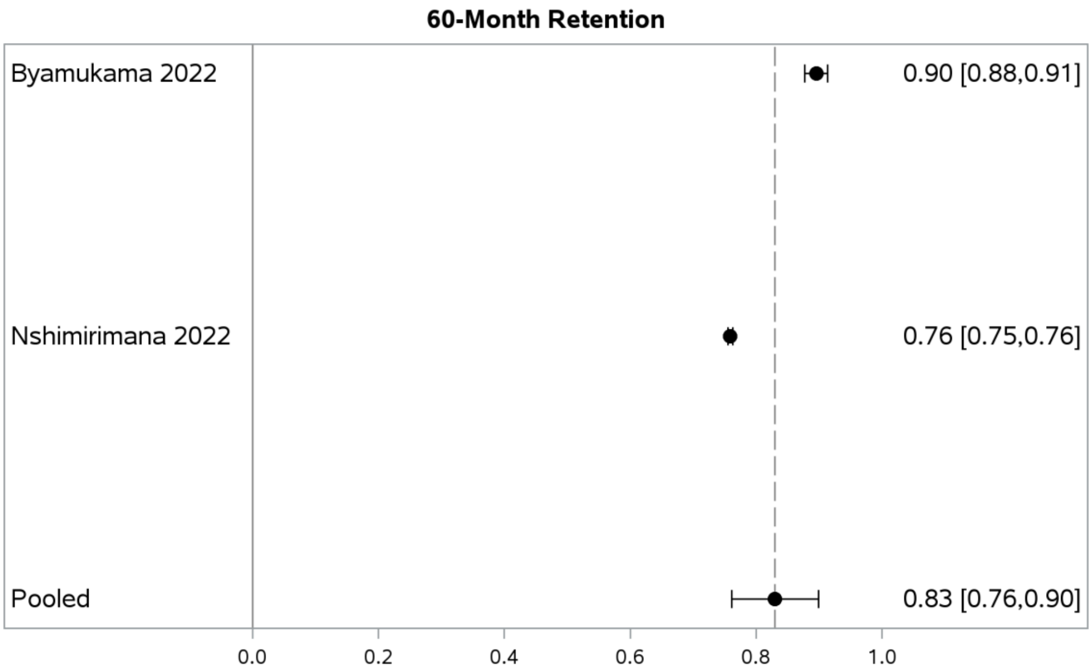


*Dashed line represents the pooled point estimate*
*Interpolated data is used for studies that did not report at that timepoint but did at the timepoint prior and after it*
**Ex. A study reported retention at 12 and 24 months therefore 18-month retention is interpolated*

**Supplemental Figure 2.** Sensitivity analysis of best case, midpoint, and worst case scenarios.

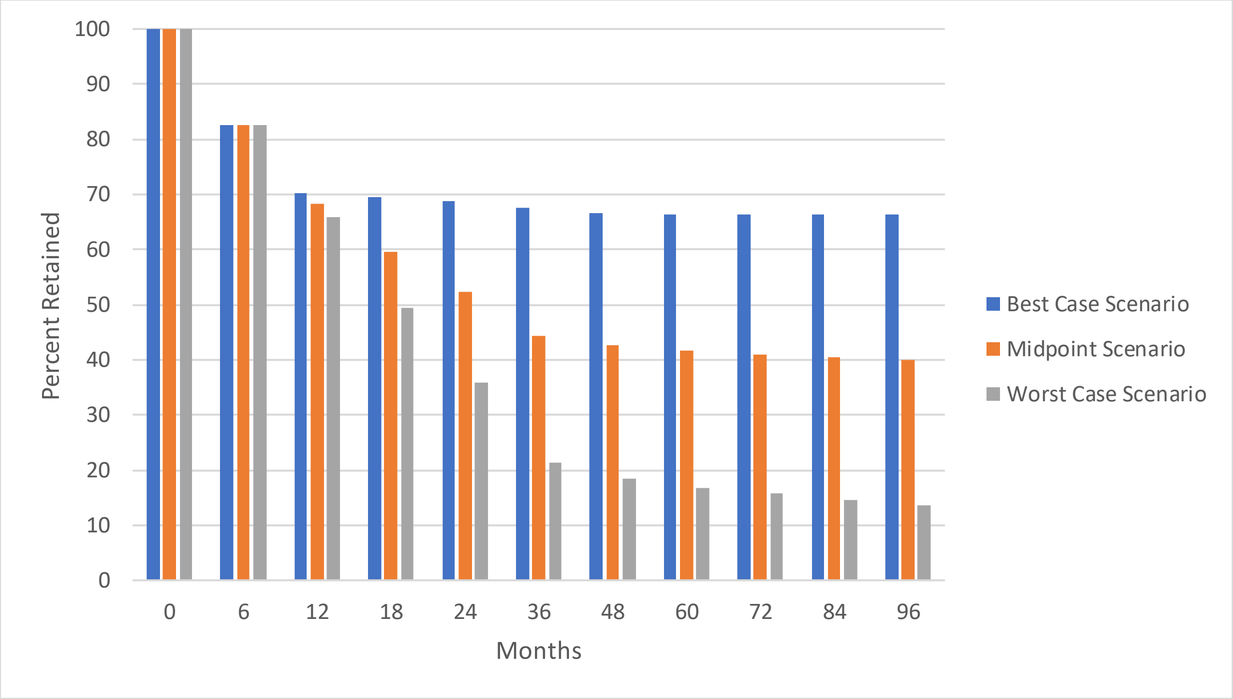

**Supplemental Table 1.** Search terms

| Database | Search terms |
|---|---|
| Pubmed 2017 – 2023 | (afghanistan[MeSH] OR "Afghanistan" OR burkina faso[MeSH] OR "burkina faso" OR burundi[MeSH] OR "Burundi" OR central african republic[MeSH] OR "central African republic" OR chad[MeSH] OR "chad" OR Democratic Republic of the Congo[Mesh] OR "Democratic Republic of Congo" OR eritrea[MeSH] OR "Eritrea" OR ethiopia[MeSH] OR "Ethiopia" OR gambia[MeSH] OR "gambia" OR guinea[MeSH] OR guinea bissau[MeSH] OR "guinea" OR "guinea bissau" OR democratic people's republic of korea[MeSH] OR republic of korea[MeSH] OR "democratic people's republic of korea" OR "republic of korea" OR liberia[MeSH] OR "Liberia" OR madagascar[MeSH] OR malawi[MeSH] OR "Madagascar" OR "Malawi" OR "Mali"[Mesh] OR "Mali" OR mozambique[MeSH] OR "Mozambique" OR niger[MeSH] OR "niger" OR rwanda[MeSH] OR "Rwanda" OR sierra leone[MeSH] OR "sierra leone" OR somalia[MeSH] OR " somalia" OR south sudan[MeSH] OR "south sudan" OR sudan[MeSH] OR "sudan" OR syria[MeSH] OR "Syria" OR "Syrian arab republic" OR togo[MeSH] OR "togo" OR uganda[MeSH] OR "Uganda" OR yemen[MeSH] OR "yemen" OR "republic of yemen" OR zambia[MeSH] OR "Zambia" OR angola[MeSH] OR "angola" OR algeria[Mesh] OR "algeria" OR bangladesh[MeSH] OR "Bangladesh" OR benin[MeSH] OR "benin" OR bhutan[MeSH] OR "Bhutan" OR bolivia[MeSH] OR "Bolivia" OR cabo verde[MeSH] OR "cabo verde" OR cambodia[MeSH] OR "Cambodia" OR cameroon[MeSH] OR "Cameroon" OR comoros[MeSH] OR "comoros" OR democratic republic of the congo[MeSH] OR "democratic republic of the congo" OR congo[MeSH] OR "congo" OR cote d'ivoire[MeSH] OR "cote d'ivoire" OR djibouti[MeSH] OR "Djibouti" OR egypt[MeSH] OR "Egypt" OR el salvador[MeSH] OR "el Salvador" OR Eswatini[MeSH] OR "eswatini" OR ghana[MeSH] OR "ghana" OR haiti[MeSH] OR "Haiti" OR honduras[MeSH] OR "Honduras" OR india[MeSH] OR "india" OR indonesia[MeSH] OR "Indonesia" OR iran[MeSH] OR "iran" OR kenya[MeSH] OR "kenya" OR Micronesia[MeSH] OR "Micronesia" OR kyrgyzstan[MeSH] OR "Kyrgyzstan" OR laos[MeSH] OR "laos" OR lebanon[MeSH] OR "Lebanon" OR lesotho[MeSH] OR "Lesotho" OR mauritania[MeSH] OR "mauritania" OR mongolia[MeSH] OR "Mongolia" OR morocco[MeSH] OR "Morocco" OR myanmar[MeSH] OR "Myanmar" OR nepal[MeSH] OR "nepal" OR nicaragua[MeSH] OR "Nicaragua" OR nigeria[MeSH] OR "Nigeria" OR pakistan[MeSH] OR "Pakistan" OR papua new guinea[MeSH] OR "papua new guinea" OR philippines[MeSH] OR "Philippines" OR samoa[MeSH] OR "samoa" OR sao tome and principe[MeSH] OR "sao tome and principe" OR senegal[MeSH] OR "senegal" OR Melanesia[Mesh] OR "Melanesia" OR sri lanka[MeSH] OR "sri lanka" OR tajikistan[MeSH] OR "Tajikistan" OR tanzania[MeSH] OR "Tanzania" OR timor leste[MeSH] OR "timor leste" OR tunisia[MeSH] OR "Tunisia" OR ukraine[MeSH] OR "Ukraine" OR uzbekistan[MeSH] OR "Uzbekistan" OR vanuatu[MeSH] OR "Vanuatu" OR vietnam[MeSH] OR "Vietnam" OR Middle East[Mesh] OR "west bank" OR "gaza" OR zimbabwe[MeSH] OR "Zimbabwe" OR albania[MeSH] OR "Albania" OR american samoa[MeSH] OR "american samoa" OR argentina[MeSH] OR "Argentina" OR armenia[MeSH] OR "Armenia" OR azerbaijan[MeSH] OR "Azerbaijan" OR republic of belarus[MeSH] OR "republic of Belarus" OR belize[MeSH] OR "Belarus" OR bosnia and herzegovina[MeSH] OR "bosnia and Herzegovina" |

| Database | Search terms |
|---|---|
| | OR botswana[MeSH] OR "Botswana" OR brazil[MeSH] OR "brazil" OR bulgaria[MeSH] OR "Bulgaria" OR china[MeSH] OR "china" OR colombia[MeSH] OR "Colombia" OR costa rica[MeSH] OR "costa rica" OR cuba[MeSH] OR "cuba" OR dominica[MeSH] OR "dominica" OR dominican republic[MeSH] OR "Dominican republic" OR ecuador[MeSH] OR "Ecuador" OR equatorial guinea[MeSH] OR "equatorial guinea" OR georgia (republic)[MeSH] OR "Georgia (republic" OR grenada[MeSH] OR "grenada" OR guatemala[MeSH] OR "Guatemala" OR guyana[MeSH] OR "Guyana" OR iraq[MeSH] OR "Iraq" OR jamaica[MeSH] OR "Jamaica" OR jordan[MeSH] OR "Jordan" OR kazakhstan[MeSH] OR "Kazakhstan" OR kosovo[MeSH] OR "Kosovo" OR libya[MeSH] OR "Libya" OR malaysia[MeSH] OR "Malaysia" OR indian ocean islands[MeSH] OR "Indian ocean islands" OR Maldives[Mesh] OR "Maldives" OR Micronesia[Mesh] OR "Micronesia" OR "marshall islands" OR Mauritius[Mesh] OR Mexico[MeSH] OR "mexico" OR "Mauritius" OR moldova[MeSH] OR "moldova" OR montenegro[MeSH] OR "Montenegro" OR Palau[MeSH] OR "palau" OR paraguay[MeSH] OR "Paraguay" OR peru[MeSH] OR "peru" OR Russia[Mesh] OR "Russia" OR "Russian Federation" OR serbia[MeSH] OR "Serbia" OR south africa[MeSH] OR "south Africa" OR saint lucia[MeSH] OR "saint lucia" OR saint vincent and the grenadines[MeSH] OR "saint vincent and the grenadines" OR suriname[MeSH] OR "suriname" OR thailand[MeSH] OR "Thailand" OR tonga[MeSH] OR "tonga" OR turkey[MeSH] OR "turkey" OR turkmenistan[MeSH] OR "Turkmenistan" OR "Tuvalu") AND (“HIV” [Mesh]) AND ("Anti-Retroviral Agents"[Mesh] OR "Agents, Anti-Retroviral" OR "Anti Retroviral Agents" OR "Antiretroviral Agents" OR "Agents, Antiretroviral" OR "Antiretroviral Agent" OR "Agent, Antiretroviral") AND ((((((("Retention in Care"[Mesh] OR "Care Retention") OR ("Continuity of Patient Care"[Mesh] OR "Care Continuity, Patient" OR "Patient Care Continuity" OR "Continuum of Care" OR "Care Continuum" OR "Continuity of Care" OR "Care Continuity" OR Retention)) OR ("Patient Dropouts"[Mesh] OR "Dropout, Patient" OR "Dropouts, Patient" OR "Patient Dropout")) OR ("Treatment Adherence and Compliance"[Mesh] OR "Therapeutic Adherence and Compliance" OR "Treatment Adherence" OR "Adherence, Treatment" OR "Therapeutic Adherence" OR "Adherence, Therapeutic")) OR ("Mortality"[Mesh] OR "Mortalities" OR "Case Fatality Rate" OR "Case Fatality Rates" OR "Rate, Case Fatality" OR "Rates, Case Fatality" OR "CFR Case Fatality Rate" OR "Crude Death Rate" OR "Crude Death Rates" OR "Death Rate, Crude" OR "Rate, Crude Death" OR "Crude Mortality Rate" OR "Crude Mortality Rates" OR "Mortality Rate, Crude" OR "Rate, Crude Mortality" OR "Death Rate" OR "Death Rates" OR "Rate, Death" OR "Mortality Rate" OR "Mortality Rates" OR "Rate, Mortality" OR "Mortality, Excess" OR "Excess Mortality" OR "Excess Mortalities" OR "Decline, Mortality" OR "Mortality Declines" OR "Mortality Decline" OR "Mortality Determinants" OR "Determinants, Mortality" OR "Determinant, Mortality" OR "Mortality Determinant" OR "Mortality, Differential" OR "Differential Mortality" OR "Differential Mortalities" OR "Age-Specific Death Rate" OR "Age-Specific Death Rates" OR "Death Rate, Age-Specific" OR "Rate, Age-Specific Death" OR "Age Specific Death Rate")) OR ("Lost to Follow-Up"[Mesh] OR "Follow-Up, Lost to" OR "Lost to Follow Up" OR "Lost to Follow-Ups")) OR ("Treatment Outcome"[Mesh] OR "Outcome, Treatment" OR "Patient-Relevant |

| Database | Search terms |
|---|---|
| | Outcome" OR "Outcome, Patient-Relevant" OR "Outcomes, Patient-Relevant" OR "Patient Relevant Outcome" OR "Patient-Relevant Outcomes") OR (nonretention[Title/Abstract]) OR (non-retention[Title/Abstract]) OR (non retention[Title/Abstract]) OR "No-Show Patients"[Mesh] OR (attendance[Title/Abstract]) OR (nonattendance[Title/Abstract]) OR (non-attendance[Title/Abstract]) OR (non attendance[Title/Abstract]) OR (Treatment interruption[Title/Abstract]) OR (Engag*[Title/Abstract]) OR (Disengag*[Title/Abstract]) OR (Reengag*[Title/Abstract]) OR (Re-engag*[Title/Abstract]) OR (Return to Care[Title/Abstract]) OR (Restart[Title/Abstract]) OR (re-enter care[Title/Abstract])) NOT (clinicaltrial[Filter]) |
| ISI Web of Science 2017 – 2023 | TS=("Anti-Retroviral Agents" OR "Agents, Anti-Retroviral" OR "Anti Retroviral Agents" OR "Antiretroviral Agents" OR "Agents, Antiretroviral" OR "Antiretroviral Agent" OR "Agent, Antiretroviral") AND TS=("Retention in Care" OR "Care Retention" OR "Continuity of Patient Care" OR "Care Continuity, Patient" OR "Patient Care Continuity" OR "Continuum of Care" OR "Care Continuum" OR "Continuity of Care" OR "Care Continuity" OR Retention OR "Patient Dropouts" OR "Dropout, Patient" OR "Dropouts, Patient" OR "Patient Dropout" OR "Treatment Adherence and Compliance" OR "Therapeutic Adherence and Compliance" OR "Treatment Adherence" OR "Adherence, Treatment" OR "Therapeutic Adherence" OR "Adherence, Therapeutic" OR "Mortality" OR "Mortalities" OR "Case Fatality Rate" OR "Case Fatality Rates" OR "Rate, Case Fatality" OR "Rates, Case Fatality" OR "CFR Case Fatality Rate" OR "Crude Death Rate" OR "Crude Death Rates" OR "Death Rate, Crude" OR "Rate, Crude Death" OR "Crude Mortality Rate" OR "Crude Mortality Rates" OR "Mortality Rate, Crude" OR "Rate, Crude Mortality" OR "Death Rate" OR "Death Rates" OR "Rate, Death" OR "Mortality Rate" OR "Mortality Rates" OR "Rate, Mortality" OR "Mortality, Excess" OR "Excess Mortality" OR "Excess Mortalities" OR "Decline, Mortality" OR "Mortality Declines" OR "Mortality Decline" OR "Mortality Determinants" OR "Determinants, Mortality" OR "Determinant, Mortality" OR "Mortality Determinant" OR "Mortality, Differential" OR "Differential Mortality" OR "Differential Mortalities" OR "Age-Specific Death Rate" OR "Age-Specific Death Rates" OR "Death Rate, Age-Specific" OR "Rate, Age-Specific Death" OR "Age Specific Death Rate" OR "Lost to Follow-Up" OR "Follow-Up, Lost to" OR "Lost to Follow Up" OR "Lost to Follow-Ups" OR "Treatment Outcome" OR "Outcome, Treatment" OR "Patient-Relevant Outcome" OR "Outcome, Patient-Relevant" OR "Outcomes, Patient-Relevant" OR "Patient Relevant Outcome" OR "Patient-Relevant Outcomes" OR "nonretention" OR "non-retention" OR "no-show patients" OR "attendance" OR "nonattendance" OR "non-attendance" OR "non attendance" OR "treatment interruption" OR "Engagement" OR "Disengagement" OR "Reengagement" OR "Re-engagement" OR "return to care" OR "Restart" OR "re-enter care")“”“”“”“”“”“”“”“”“”“”“”“”“”“”“”“”“”“”“”“”“”“”“”“”“”“”“”“”“”“”“”“”“”“”“”“”“”“”“”“”“”“”“”“”“”“”“ ”“”“”“”“”“”“”“”“”“”“”“”“”“”AND TS=(afghanistan OR Afghanistan OR "burkina faso" OR "burkina faso" OR burundi OR Burundi OR "central african republic" OR "central African republic" OR chad OR chad OR "Democratic Republic of the Congo" OR "Democratic Republic of Congo" OR eritrea OR Eritrea OR ethiopia OR Ethiopia OR gambia OR gambia OR |

| Database | Search terms |
| --- | --- |
|  | guinea OR "guinea bissau" OR guinea OR "guinea bissau" OR "democratic people's republic of korea" OR "republic of korea" OR "democratic people's republic of korea" OR "republic of korea" OR liberia OR Liberia OR madagascar OR malawi OR Madagascar OR Malawi OR Mali OR Mali OR mozambique OR Mozambique OR niger OR niger OR rwanda OR Rwanda OR "sierra leone" OR "sierra leone" OR somalia OR " somalia" OR "south sudan" OR "south sudan" OR sudan OR sudan OR syria OR Syria OR "Syrian arab republic" OR togo OR togo OR uganda OR Uganda OR yemen OR yemen OR "republic of yemen" OR zambia OR Zambia OR angola OR angola OR algeria OR algeria OR bangladesh OR Bangladesh OR benin OR benin OR bhutan OR Bhutan OR bolivia OR Bolivia OR "cabo verde" OR "cabo verde" OR cambodia OR Cambodia OR cameroon OR Cameroon OR comoros OR comoros OR "democratic republic of the congo" OR "democratic republic of the congo" OR congo OR congo OR "cote d'ivoire" OR "cote d'ivoire" OR djibouti OR Djibouti OR egypt OR Egypt OR "el salvador" OR "el Salvador" OR Eswatini OR eswantini OR ghana OR ghana OR haiti OR Haiti OR honduras OR Honduras OR india OR india OR indonesia OR Indonesia OR iran OR iran OR kenya OR kenya OR Micronesia OR Micronesia OR kyrgyzstan OR Kyrgyzstan OR laos OR laos OR lebanon OR Lebanon OR lesotho OR Lesotho OR mauritania OR mauritania OR mongolia OR Mongolia OR morocco OR Morocco OR myanmar OR Myanmar OR nepal OR nepal OR nicaragua OR Nicaragua OR nigeria OR Nigeria OR pakistan OR Pakistan OR "papua new guinea" OR "papua new guinea" OR philippines OR Philippines OR samoa OR samoa OR "sao tome" AND principe OR "sao tome and principe" OR senegal OR senegal OR Melanesia OR Melanesia OR "sri lanka" OR "sri lanka" OR tajikistan OR Tajikistan OR tanzania OR Tanzania OR "timor leste" OR "timor leste" OR tunisia OR Tunisia OR ukraine OR Ukraine OR uzbekistan OR Uzbekistan OR vanuatu OR Vanuatu OR vietnam OR Vietnam OR "Middle East" OR "west bank" OR gaza OR zimbabwe OR Zimbabwe OR albania OR Albania OR "american samoa" OR "american samoa" OR argentina OR Argentina OR armenia OR Armenia OR azerbaijan OR Azerbaijan OR "republic of belarus" OR "republic of Belarus" OR belize OR Belarus OR bosnia AND herzegovina OR "bosnia and Herzegovina" OR botswana OR Botswana OR brazil OR brazil OR bulgaria OR Bulgaria OR china OR china OR colombia OR Colombia OR "costa rica" OR "costa rica" OR cuba OR cuba OR dominica OR dominica OR "dominican republic" OR "Dominican republic" OR ecuador OR Ecuador OR "equatorial guinea" OR "equatorial guinea" OR georgia (republic) OR "Georgia (republic" OR grenada OR grenada OR guatemala OR Guatemala OR guyana OR Guyana OR iraq OR Iraq OR jamaica OR Jamaica OR jordan OR Jordan OR kazakhstan OR Kazakhstan OR kosovo OR Kosovo OR libya OR Libya OR malaysia OR Malaysia OR "indian ocean islands" OR "Indian ocean islands" OR Maldives OR Maldives OR Micronesia OR Micronesia OR "marshall islands" OR Mauritius OR Mexico OR mexico OR Mauritius OR moldova OR moldova OR montenegro OR Montenegro OR Palau OR palau OR paraguay OR Paraguay OR peru OR peru OR Russia OR Russia OR "Russian Federation" OR serbia OR Serbia OR "south africa" OR "south Africa" OR "saint lucia" OR "saint lucia" OR "saint vincent" AND "the grenadines" OR "saint vincent and the grenadines" OR suriname OR suriname OR thailand OR Thailand OR tonga OR tonga OR turkey OR turkey OR turkmenistan OR Turkmenistan OR Tuvalu) |

| Database | Search terms |
|---|---|
| Cochrane Database of Systematic Reviews (up to 2023) | ((MeSH descriptor: [Anti-Retroviral Agents] explode all trees) OR ("Agents, Anti-Retroviral" OR "Anti Retroviral Agents" OR "Antiretroviral Agents" OR "Agents, Antiretroviral" OR "Antiretroviral Agent" OR "Agent, Antiretroviral")) AND ((MeSH descriptor: [Retention in Care] explode all trees) OR ("Care Continuity, Patient" OR "Patient Care Continuity" OR "Continuum of Care" OR "Care Continuum" OR "Continuity of Care" OR "Care Continuity" OR Retention)) OR ((MeSH descriptor: [Patient Dropouts] explode all trees) OR ("Dropout, Patient" OR "Dropouts, Patient" OR "Patient Dropout")) OR ((MeSH descriptor: [Treatment Adherence and Compliance] explode all trees) OR ("Therapeutic Adherence and Compliance" OR "Treatment Adherence" OR "Adherence, Treatment" OR "Therapeutic Adherence" OR "Adherence, Therapeutic")) OR ((MeSH descriptor: [Mortality] explode all trees) OR ("Mortalities" OR "Case Fatality Rate" OR "Case Fatality Rates" OR "Rate, Case Fatality" OR "Rates, Case Fatality" OR "CFR Case Fatality Rate" OR "Crude Death Rate" OR "Crude Death Rates" OR "Death Rate, Crude" OR "Rate, Crude Death" OR "Crude Mortality Rate" OR "Crude Mortality Rates" OR "Mortality Rate, Crude" OR "Rate, Crude Mortality" OR "Death Rate" OR "Death Rates" OR "Rate, Death" OR "Mortality Rate" OR "Mortality Rates" OR "Rate, Mortality" OR "Mortality, Excess" OR "Excess Mortality" OR "Excess Mortalities" OR "Decline, Mortality" OR "Mortality Declines" OR "Mortality Decline" OR "Mortality Determinants" OR "Determinants, Mortality" OR "Determinant, Mortality" OR "Mortality Determinant" OR "Mortality, Differential" OR "Differential Mortality" OR "Differential Mortalities" OR "Age-Specific Death Rate" OR "Age-Specific Death Rates" OR "Death Rate, Age-Specific" OR "Rate, Age-Specific Death" OR "Age Specific Death Rate")) OR ((MeSH descriptor: [Lost to Follow-Up] explode all trees) OR (“Follow-Up, Lost to" OR "Lost to Follow Up" OR "Lost to Follow-Ups")) OR ((MeSH descriptor: [Treatment Outcome] explode all trees) OR ("Outcome, Treatment" OR "Patient-Relevant Outcome" OR "Outcome, Patient-Relevant" OR "Outcomes, Patient-Relevant" OR "Patient Relevant Outcome" OR "Patient-Relevant Outcomes") OR ("nonretention" OR "non-retention" OR "non retention" OR "attendance" OR "nonattendance" OR "non-attendance" OR "non attendance" OR "treatment interruption" OR "engagement" OR "disengagement" OR "reengagement" OR "re-engagement" OR "return to care" or "restart" OR "re-enter care") OR MeSH descriptor: [No-Show Patients] explode all trees) AND (Afghanistan OR Burkina Faso OR Burundi OR Central African Republic OR Chad OR Democratic Republic of Congo OR Eritrea OR Ethiopia OR The Gambia OR Guinea OR Guinea-Bissau OR Democratic People’s Republic of Korea OR Liberia OR Madagascar OR Malawi OR Mali OR Mozambique OR Niger OR Rwanda OR Sierra Leone OR Somalia OR South Sudan OR Sudan OR Syrian Arab Republic OR Togo OR Uganda OR Republic of Yemen OR Zambia OR Angola OR Algeria OR Bangladesh OR Benin OR Bhutan OR Bolivia OR Cabo Verde OR Cambodia OR Cameroon OR Comoros OR Republic of Congo OR Ivory Coast OR Djibouti OR Egypt OR El Salvador OR Eswatini OR Ghana OR Haiti OR Honduras OR India OR Indonesia OR Iran OR Kenya OR Kiribati OR Kyrgyzstan OR Laos OR Lebanon OR Lesotho OR Mauritania OR Micronesia OR Mongolia OR Morocco OR Myanmar OR Nepal OR Nicaragua OR Nigeria OR Pakistan OR Papua New Guinea OR Philippines OR Samoa OR Sao Tome and Principe OR Senegal OR Solomon Islands OR |

| Database | Search terms |
|---|---|
| | Sri Lanka OR Tanzania OR Tajikistan OR Timor OR Tunisia OR Ukraine OR Uzbekistan OR Vanuatu OR Vietnam OR West Bank and Gaza OR Zimbabwe OR Albania OR American Samoa OR Argentina OR Armenia OR Azerbaijan OR Belarus OR Belize OR Bosnia and Herzegovina OR Botswana OR Brazil OR Bulgaria OR China OR Colombia OR Costa Rica OR Cuba OR Dominica OR Dominican Republic OR Ecuador OR Equatorial Guinea OR Georgia OR Grenada OR Guatemala OR Guyana OR Iraq OR Jamaica OR Jordan OR Kazakhstan OR Kosovo OR Libya OR Malaysia OR Maldives OR Marshall Islands OR Mauritius OR Mexico OR Moldova OR Montenegro OR Palau OR Paraguay OR Peru OR Russian Federation OR Serbia OR South Africa OR Saint Lucia OR Saint Vincent and the Grenadines OR Suriname OR Thailand OR Tonga OR Turkey OR Turkmenistan OR Tuvalu) |
| EMBASE 2017 - 2023 | ('afghanistan'/exp OR afghanistan OR 'burkina faso'/exp OR 'burkina faso' OR 'burundi'/exp OR burundi OR 'central african republic'/exp OR 'central african republic' OR 'chad'/exp OR chad OR 'democratic republic of congo' OR 'eritrea'/exp OR eritrea OR 'ethiopia'/exp OR ethiopia OR 'gambia'/exp OR gambia OR 'guinea'/exp OR 'guinea bissau'/exp OR guinea OR 'guinea bissau' OR 'republic of korea'/exp OR 'democratic peoples republic of korea' OR 'republic of korea' OR 'liberia'/exp OR liberia OR 'madagascar'/exp OR 'malawi'/exp OR madagascar OR malawi OR 'mali'/exp OR mali OR 'mozambique'/exp OR mozambique OR 'niger'/exp OR niger OR 'rwanda'/exp OR rwanda OR 'sierra leone'/exp OR 'sierra leone' OR 'somalia'/exp OR 'somalia' OR 'south sudan'/exp OR 'south sudan' OR 'sudan'/exp OR sudan OR 'syria'/exp OR syria OR 'syrian arab republic' OR 'togo'/exp OR togo OR 'uganda'/exp OR uganda OR 'yemen'/exp OR yemen OR 'republic of yemen' OR 'zambia'/exp OR zambia OR 'angola'/exp OR angola OR 'algeria'/exp OR algeria OR 'bangladesh'/exp OR bangladesh OR 'benin'/exp OR benin OR 'bhutan'/exp OR bhutan OR 'bolivia'/exp OR bolivia OR 'cabo verde'/exp OR 'cabo verde' OR 'cambodia'/exp OR cambodia OR 'cameroon'/exp OR cameroon OR 'comoros'/exp OR comoros OR 'democratic republic of the congo'/exp OR 'democratic republic of the congo' OR 'congo'/exp OR congo OR 'cote d ivoire' OR 'djibouti'/exp OR djibouti OR 'egypt'/exp OR egypt OR 'el salvador'/exp OR 'el salvador' OR eswatini OR eswantini OR 'ghana'/exp OR ghana OR 'haiti'/exp OR haiti OR 'honduras'/exp OR honduras OR 'india'/exp OR india OR 'indonesia'/exp OR indonesia OR 'iran'/exp OR iran OR 'kenya'/exp OR kenya OR 'micronesia'/exp OR micronesia OR 'kyrgyzstan'/exp OR kyrgyzstan OR 'laos'/exp OR laos OR 'lebanon'/exp OR lebanon OR 'lesotho'/exp OR lesotho OR 'mauritania'/exp OR mauritania OR 'mongolia'/exp OR mongolia OR 'morocco'/exp OR morocco OR 'myanmar'/exp OR myanmar OR 'nepal'/exp OR nepal OR 'nicaragua'/exp OR nicaragua OR 'nigeria'/exp OR nigeria OR 'pakistan'/exp OR pakistan OR 'papua new guinea'/exp OR 'papua new guinea' OR 'philippines'/exp OR philippines OR 'samoa'/exp OR samoa OR 'sao tome and principe'/exp OR 'sao tome and principe' OR 'senegal'/exp OR senegal OR 'melanesia'/exp OR melanesia OR 'sri lanka'/exp OR 'sri lanka' OR 'tajikistan'/exp OR tajikistan OR 'tanzania'/exp OR tanzania OR 'timor leste'/exp OR 'timor leste' OR 'tunisia'/exp OR tunisia OR 'ukraine'/exp OR ukraine OR 'uzbekistan'/exp OR uzbekistan OR 'vanuatu'/exp OR vanuatu OR 'vietnam'/exp OR vietnam OR 'middle east'/exp OR 'west bank' OR gaza OR 'zimbabwe'/exp OR zimbabwe OR |

| Database | Search terms |
|---|---|
| | 'albania'/exp OR albania OR 'american samoa'/exp OR 'american samoa' OR 'argentina'/exp OR argentina OR 'armenia'/exp OR armenia OR 'azerbaijan'/exp OR azerbaijan OR 'republic of belarus'/exp OR 'republic of belarus' OR 'belize'/exp OR belarus OR 'bosnia and herzegovina'/exp OR 'bosnia herzegovina' OR 'bosnia and herzegovina' OR 'bosnia and herzegowina' OR 'bosnia-herzegovina' OR 'botswana'/exp OR botswana OR 'brazil'/exp OR brazil OR 'bulgaria'/exp OR bulgaria OR 'china'/exp OR china OR 'colombia'/exp OR colombia OR 'costa rica'/exp OR 'costa rica' OR 'cuba'/exp OR cuba OR 'dominica'/exp OR dominica OR 'dominican republic'/exp OR 'dominican republic' OR 'ecuador'/exp OR ecuador OR 'equatorial guinea'/exp OR 'equatorial guinea' OR 'georgia (republic)'/exp OR 'georgia (republic)' OR 'georgian s.s.r.' OR 'georgian ssr' OR 'georgian soviet socialist republic' OR 'grenada'/exp OR grenada OR 'guatemala'/exp OR guatemala OR 'guyana'/exp OR guyana OR 'iraq'/exp OR iraq OR 'jamaica'/exp OR jamaica OR 'jordan'/exp OR jordan OR 'kazakhstan'/exp OR kazakhstan OR 'kosovo'/exp OR kosovo OR 'libya'/exp OR libya OR 'malaysia'/exp OR malaysia OR 'indian ocean islands'/exp OR 'indian ocean islands' OR 'maldives'/exp OR maldives OR 'micronesia'/exp OR micronesia OR 'marshall islands' OR 'mauritius'/exp OR 'mexico'/exp OR mexico OR mauritius OR 'moldova'/exp OR moldova OR 'montenegro'/exp OR montenegro OR 'palau'/exp OR palau OR 'paraguay'/exp OR paraguay OR 'peru'/exp OR peru OR 'russia'/exp OR russia OR 'russian federation' OR 'serbia'/exp OR serbia OR 'south africa'/exp OR 'south africa' OR 'saint lucia'/exp OR 'saint lucia' OR 'saint vincent and the grenadines'/exp OR 'saint vincent and the grenadines' OR 'st. vincent and the grenadines' OR 'suriname'/exp OR suriname OR 'thailand'/exp OR thailand OR 'tonga'/exp OR tonga OR 'turkey'/exp OR turkey OR 'turkmenistan'/exp OR turkmenistan OR Tuvalu) AND ('antiretrovirus agent'/exp OR 'anti retroviral agent' OR 'anti retroviral agents' OR 'anti-retroviral agents' OR 'antiretroviral agent' OR 'antiretroviral agents' OR 'antiretrovirus agent') AND (('retention in care'/exp OR 'care retention' OR 'retention in care') OR ('patient care'/exp OR 'care, continuity of' OR 'continuity of care' OR 'continuity of patient care' OR 'episode of care' OR 'patient care' OR 'patient care management' OR 'patient care team' OR 'patient centered care' OR 'patient helper' OR 'patient management' OR 'patient navigation' OR 'patient-centered care') OR ('patient dropout'/exp OR 'care drop out' OR 'care dropout' OR 'dropout in inpatients' OR 'dropout in outpatients' OR 'dropout in patients' OR 'outpatient dropout' OR 'patient drop out' OR 'patient drop outs' OR 'patient dropout' OR 'patient dropouts' OR 'therapy drop out' OR 'therapy dropout' OR 'treatment drop out' OR 'treatment dropout') OR ('patient compliance'/exp OR 'adherence to therapy' OR 'adherence to treatment' OR 'compliance to therapy' OR 'compliance to treatment' OR 'patient adherence' OR 'patient compliance' OR 'patients` adherence' OR 'therapy adherence' OR 'therapy compliance' OR 'treatment adherence' OR 'treatment adherence and compliance' OR 'treatment compliance') OR ('mortality'/exp OR 'mortality' OR 'mortality model') OR ('follow up'/exp OR 'follow up' OR 'follow up study' OR 'follow-up studies' OR 'followup' OR 'lost to follow up' OR 'lost to follow-up') OR ('treatment outcome'/exp OR 'patient outcome' OR 'therapeutic outcome' OR 'therapy outcome' OR 'treatment outcome') OR "nonretention" OR "non-retention" OR "no-show patients" OR "attendance" OR |

| Database | Search terms |
|---|---|
| | "nonattendance" OR "non-attendance" OR "non attendance" OR "treatment interruption" OR "Engagement" OR "Disengagement" OR "Reengagement" OR "Re-engagement" OR "return to care" OR "Restart" OR "re-enter care") NOT ('randomized controlled trial'/exp OR 'controlled trial, randomized' OR 'randomized controlled trial' OR 'randomised controlled trial' OR 'randomized controlled study' OR 'randomized controlled trial' OR 'trial, randomized controlled' OR ‘case report’) |
| IAS abstract archives 2017, 2019, 2021, 2023 | retention OR attrition OR “loss to follow up” OR "lost to follow-up" OR "loss to follow up" OR "loss to follow-up" |
| International AIDS conference abstract archives 2016, 2018, 2020, 2022 | retention OR attrition OR "lost to follow up" OR "lost to follow-up" OR "loss to follow up" OR "loss to follow-up" |
| CROI abstract archives 2017 – 2023 | retention OR attrition OR "lost to follow up" OR "lost to follow-up" OR "loss to follow up" OR "loss to follow-up" |

**Supplemental Table 2.** Characteristics of studies included in the systematic review

| Reference | Region | Facilities (N) | Sector | UTT Implementation Year | Dates of Cohort Enrollment | Dates of Analytical Cohort Enrollment | Analytical Cohort (N) | Median Age (IQR) | Female (%) | Median CD4 Count (IQR) | Key Populations | Notes |
|---|---|---|---|---|---|---|---|---|---|---|---|---|
| Ahmed 2020 | Sub Saharan Africa | 11 health facilities (eight health centers and three hospitals) that had > 20 PLHIV on ART by the end of December 2018 were included in the study. | Urban | 2016 | October 20 2016 - July 18 2018 | October 20 2016 - July 18 2018 | 988 | 33 (27-40) | 52.6 | - | No | |
| Alhaj 2019 | Sub Saharan Africa | | Urban; Rural | 2016 | June 2015 and August 2016 | June 2015 and August 2016 | 1492 | 33 (26-41) | 62.5 | - | No | Includes 20 people who are aged 10-19 |
| Atuhaire 2022 | Sub Saharan Africa | | Urban | 2016 | January 2018 - December 2020 | January 2018 - December 2020 | 275 | - | 100 | - | Female Sex Workers | |
| Avalos 2019 | Sub Saharan Africa | | Urban; Rural | 2016 | | | 1523 | 39 (32-48) | 63 | - | No | Age and Female are reported on the full cohort not the analytical cohort |
| Benzekri 2022 | West Africa | Services des Maladies Infectieuses et Tropicales, CHNU-Fann in Dakar and the Centre de SantÃ© de Ziguinchor | Urban | 2016 | April 2017 - April 2018 | April 2017 - April 2018 | 207 | 37 (31-46) | 69.6 | 181 (63-382) | No | They included people who had HIV-2/HIV-1 and 2 so we'll also need to note that when we do our sensitivity analysis |
| Byamukama 2022 | Sub Saharan Africa | Kabwohe has a catchment area of 15 districts that have a combined | Rural | 2016 | January 2016 - December 2020 | January 2016 - December 2020 | 976 | 35 | 57.1 | - | No | Mean follow-up period is reported |

| | | | | | | | | | | | | |
|---|---|---|---|---|---|---|---|---|---|---|---|---|
| | | population of 8 874 862 people<br>5500 PLHIV in active care | | | | | | | | | | |
| Cassidy 2023 | Sub Saharan Africa | | Other | 2016 | January 1 2014 - April 1 2018 | September 2 2016 - April 1 2018 | 1553 | Male clinics: 31.2 (26.9-36)<br>General clinics: 31.3 (27.1-36.2) | 0 | Male clinics: 343 (233-455)<br>General clinics: 335 (193-484) | No | Median age and CD4 includes the full cohort (i.e., people who enrolled in 2014) and is based on imputation and propensity score matching values<br>Data extracted only includes the cohort who initiated after September 1, 2016 and is from the primary tables. The date of UTT as the tables provided did not allow us to calculate it from 2015 onwards by year.<br>Based on the attrition definition, attrition is equivalent to LTFU. |
| Chauke 2020 | Sub Saharan Africa | | Urban | 2016 | November 2016 | November 2016 | 367 | 36.3 (9.6) | 63 | - | No | |
| Dorward 2020 | Sub Saharan Africa | | Urban; Rural | 2016 | September 1 2016 - August 31 2017 | September 1 2016 - August 31 2017 | 4952 | 32.4 (27.2-39.7) | 59.9 | - | No | Mean follow-up period is reported |
| Dumchev 2022 | Eastern Europe | 50 | Urban; Rural | 2015 | October 1 2017 - June 30 2018 | October 1 2017 - June 30 2018 | 1057 | 37 (16-76) | 51.7 | - | No | |
| Eamsakulrat 2022 | Southeast Asia | 1200 bed | Urban | 2014 | January 1 2015 - December 31 2017 | January 1, 2015 - December 31, 2017 | 270 | 36.6 (18.3-79.8) | 21.8 | 159 (53-308) | No | Age and CD4 is on full cohort |
| Ntamatungiro 2021 | Sub Saharan Africa | | Rural | 2016 | December 16 2016 - December 15 2017 | December 16 2016 - December 15 2017<br>March 16 2019 - | 902 | 38 (30-45) | 64.3 | - | No | |

| | | | | | | | | | | | |
|---|---|---|---|---|---|---|---|---|---|---|---|
| | | | | | March 16 2019 - September 15 2020 | September 15 2020 | | | | | |
| Eshiwani 2018 | Sub Saharan Africa | | Urban | 2016 | September 2016 - June 2017 | September 2016 - June 2017 | 167 | - | - | - | No | A subset of the entire cohort was used as they did not explicitly report death or transfers on the delayed treatment arm. |
| Harooni 2022 | Central Asia | | Urban; Rural | 2016 | 2018 | 2018 | 124 | - | 25.2 | - | No | Female includes the entire cohort |
| Hirasen 2020 | Sub Saharan Africa | | Urban | 2016 | December 1 2016 - May 31 2017 | December 1 2016 - May 31 2017 | 1143 | 36.6 (30.6 - 43.9) | 58.8 | 224 (93-369) | No | Age at enrollment and Median CD4 is based on full cohort |
| Hoang 2022 | Southeast Asia | | Urban; Rural | 2015 | January 1 2011 - December 31 2016 | January 1 2015 - December 31 2015 | 4892 | - | - | - | No | |
| Ibiloye 2018 | Sub Saharan Africa | | Urban | 2016 | August 1 2016 - February 28 2017 | August 1 2016 - February 28 2017 | 710 | 30 (24-35) | 77.3 | - | Commercial Sex Workers, MSM, PWID + Partners | |
| Ibiloye 2021 | Sub Saharan Africa | | Urban; Rural | 2016 | January 1 2016 - December 21 2019 | January 1 2016 - December 21 2019 | 3495 | 34 (29-40) | 60.6 | - | MSM, FSW, PWID, Transgender people | |
| Izudi 2022 | Sub Saharan Africa | 20,000 - 100,000 | Urban | 2016 | March 1, 2018 - February 28, 2020 | March 1 2018 - February 28 2020 | 9952 | 32.7 (8.8) | 66.3 | - | No | |
| Jamieson 2021 | Sub Saharan Africa | | Urban; Rural | 2017 | January 1 2019 - December 31 2020 | January 1 2019 - December 31 2020 | 32197 | 37 (29-44) | 60.8 | - | No | |
| Januraga 2018 | Southeast Asia | | Urban | 2013 | September 15 2015 - September 30 2016 | September 15 2015 - September 30 2016 | 606 | 27.3 (23.1-32.9) | 15 | - | MSM, FSW, PWID, Transgender women | Age and Female are based on full cohort |
| Johansson 2021 | Sub Saharan Africa | | Urban | 2016 | June 2015 - September 2018 | June 2015 - September 2018 | 2043 | 32 (27-40) | 59 | - | No | Age and female is on full cohort |
| Kimanga 2022 | Sub Saharan africa | | Urban; Rural | 2016 | July 2015 - June 2018 | July 2015 - June 2018 | 8592 | 35.1 (28.5-43.5) | 68.2 | - | No | |
| Lilian 2020 | Sub Saharan Africa | | Urban; Rural | 2016 | October 2017 - June 2018 | October 2017 - June 2018 | 32290 | Same day: 31.9 (15.1- | 65.2 | - | No | |

| | | | | | | | | | | | | |
|---|---|---|---|---|---|---|---|---|---|---|---|---|
| | | | | | | | | 79.6)<br>1-7 days: 34.8 (15.3-75.7)<br>8-21 days: 35.8 (15.0-79.4)<br>22+ days: 35.4 (15.0-78.2) | | | | |
| Makurumidze 2020 | Sub Saharan Africa | | Urban; Rural | 2016 | April 2016 - May 2016<br>January 2017 - February 2017 | April 2016 - May 2016<br>January 2017 - February 2017 | 3636 | Before: 37 (30-44)<br>After: 36 (29-43) | 62.1 | - | No | Age, Median Follow-up time, and Female includes full cohort |
| Matare 2020 | Sub Saharan Africa | 52% of all people on ART in Harare province | Urban | 2016 | Before Treat-All: April 2015 - June 2015<br>Post Treat-All: April 2017 - June 2017 | Before Treat-All: April 2015 - June 2015<br>Post Treat-All: April 2017 - June 2017 | 3971 | 31 (27-41) | 58.5 | - | No | |
| Matthews 2020 | Sub Saharan Africa | | Urban; Rural | 2016 | March 2015 - September 2016 | March 2015 - September 2016 | 437 | Uganda: 29 (9)<br>South Africa: 31 (9) | 100 | Uganda: 477 (421-618)<br>South Africa: 440 (394-490) | Pregnant and Postpartum Women | Weighted average for age was calculated |
| Mayasi 2022 | Sub Saharan Africa | <100 - >500 | Urban | 2016 | January 1 2010 - December 31 2019 | November 1 2016 - December 31 2019 | 11281 | 40.1 (11.5) | 66 | 296 (187-483) | No | |
| Mshweshwe-Pakela 2020 | Sub Saharan Africa | | Urban | 2016 | January 1 2017 - July 31 2017 | January 1 2017 - July 31 2017 | 710 | 32 (27-39) | 65.2 | 312 (156-490) | No | Age and CD4 is based on full cohort |
| Nshimirimana 2022 | Sub Saharan Africa | | Urban; Rural | 2016 | January 1 2015 - July 31 2020 | January 1 2015 - July 31 2020 | 29829 | 34 (26-43) | 68.6 | - | No | |

| | | | | | | | | | | | |
|---|---|---|---|---|---|---|---|---|---|---|---|
| Onoya 2021 | Sub Saharan Africa | | Urban | 2016 | April 2015 - December 2015 (Pre UTT)<br>July 2017 - August 2017 (UTT)<br>October 2017 - August 2018 (SDI) | April 2015 - November 2015<br>May 2017 - September 2017 | 297 | 33.5 (28.3-39.4) | 56.1 | 237 (100-430) | No | |
| Opio 2019 | Sub Saharan Africa | | Urban | 2016 | January 1 2015 - December 31 2017 | January 1 2015 - December 31 2017 | 646 | - | 60.5 | - | No | Weighted average for median follow-up time could not be calculated as the Ns by enrollment period are not provided, so a mean of the two median follow-up times was taken for the purposes of this analysis |
| Opito 2020 | Sub Saharan Africa | 8066 | Urban; Rural | 2016 | June 2017 - May 2018 | June 2017 - May 2018 | 536 | 38.0 (13.1) | 56.5 | - | No | Includes 37 patients <20 years of age |
| Pillay 2019 | Sub Saharan Africa | | Urban | 2016 | January 2016 - December 2016 | January 2016 - December 2016 | 120 | 39 (12) | 55 | 228 | No | |
| Rogers 2021 | Sub Saharan Africa | 3500-5000 | Urban | 2016 | July 2016 - September 2016 | July 2016 - September 2016 | 423 | 39.72 (8) | 65.08 | 225 (101-411) | No | |
| Romo 2022 | Sub Saharan Africa | | Urban; Rural | 2016 | March 2015 - October 2020 | March 2015 - October 2020 | 3563 | - | 47.2 | 135 (50-309) | No | Database closure varied due to multiple sites across various countries |
| Ross 2019 | Central Africa | | Urban; Rural | 2016 | July 1 2014 - September 13 2017 | July 2016 - September 2017 | 1082 | 33 (27-39) | 56.8 | 392 (219-586) | No | |
| Seekaew 2019 | Southeast Asia | | Urban | 2017 | July 2017 - April 2019 | July 2017 - April 2019 | 180 | 26.4 (23.3-30.0) | - | 306 (232-442) | Transgender Women | Retention Ns were taken from the text not the table.<br>The age and CD4 were taken from the table which a subset of the analytic cohort.<br>Includes people < 25 but based on the table the minimum age was 17.6 |

| | | | | | | | | | | | | |
|---|---|---|---|---|---|---|---|---|---|---|---|---|
| Seekaew 2021 | Southeast Asia | | Urban | 2017 | July 2017 - July 2018 | July 2017 - July 2018 | 1904 | 28.3 (23.8-35.1) | - | 294 (192-415) | Includes MSM and TGW | |
| Ssempijja 2020 | Sub Saharan Africa | | Urban | 2016 | 2006 -2018 | 2015 - 2016 | 1305 | - | 71 | 302 (113-476) | No | |
| Stafford 2019 | Sub Saharan Africa | | Urban; Rural | 2016 | October 2015 - September 2016 | October 2015 - September 2016 | 2652 | 33 (27-40) | 67 | 323 (161-518) | No | Age and CD4 are based on full cohort |
| Stockton 2021 | Sub Saharan Africa | | Urban | 2016 | April 2017 - October 2017 (Clinic A)<br>April 2017 - March 2018 (Clinic B) | April 2017 - October 2017 (Clinic A)<br>April 2017 - March 2018 (Clinic B) | 1091 | 33.5 (9.6) | 53 | - | No | Retained at 6-months is based on "In care and after 6 months " definition<br><br>Alive and on ART is based on the "Currently on ART" definition |
| Teshale 2020 | Sub Saharan Africa | 5573 | Urban | 2016 | January 1 2015 - December 31 2018 | January 1 2015 - December 31 2018 | 531 | 32 (25-40) | 57.4 | 250 (110-429) | No | |
| Bernard Marc 2023 | Caribbean | | Urban | 2016 | December 2020 - June 2022 | December 2020 - June 2022 | 193 | 40 (33-47) | 47 | - | No | |
| Chagomerana 2023 | Sub Saharan Africa | | Urban; Rural | 2016 | May 2015 - June 2016 | May 2015 - June 2016 | 291 | 27 (23-30) | 100 | - | No | N for the analytical cohort excludes 8 people who withdrew consent, but demographic characteristics includes the 8 who withdrew consent |
| Dorward 2023 | Sub Saharan Africa | | Urban; Rural | 2016 | December 1, 2019 - December 1, 2020 | December 1, 2019 - December 1, 2020 | 22821 | 31 (26.0-38.0) | 63.5 | - | No | Median follow-up time is based on full cohort which includes the analysis comparing those who are treatment-experienced |
| Gemechu 2023 | Sub Saharan Africa | | 0 | 2016 | October 2020 - July 2021 | October 2020 - July 2021 | 235 | 33.9 (12.1) | 70.6 | - | No | |
| Govere 2023 | Sub Saharan Africa | 383869 | Urban; Rural | 0 | June 2020 - November 2020 | June 2020 - November 2020 | 403 | 0 | 45.2 | - | No | |
| Hamooya 2023 | Sub Saharan Africa | | Urban; Rural | 2016 | January 1, 2014 - October 1, 2020 | August 1, 2016 - October 1, 2020 | 3649 | 37 (30-45) | 59.8 | - | No | |

| | | | | | | | | | | | | |
|---|---|---|---|---|---|---|---|---|---|---|---|---|
| Joaquim 2023 | Sub Saharan Africa | 16244 | Urban | 2016 | January 1, 2016 - December 30, 2018 | January 1, 2016 - December 30, 2018 | 1247 | 42 (18-88) | 50.5 | 359 (4.0 - 1765.0) | No | |
| Kimanga 2023 | Sub Saharan Africa | | Urban; Rural | 0 | April 2018 - March 2021 | April 2018 - March 2021 | 7046 | 0 | 66.7 | - | No | |
| Lavoie 2023 | Sub Saharan Africa | | Urban; Rural | 2016 | April 2018 - March 2019 | April 2018 - March 2019 | 75348 | 0 | 69.6 | - | No | |
| MacKellar 2022 | Sub Saharan Africa | | Urban; Rural | 2016 | March 1, 2016 - March 31, 2018 | March 1, 2016 - March 31, 2018 | 769 | 0 | 58.5 | - | No | |
| Masaba 2023 | Sub Saharan Africa | | Urban | 2016 | October 2016 - September 2019 | October 2016 - September 2019 | 1515 | 46.4 (39.6 - 54.6) | 56.4 | 159 (83-273) | No | Mean follow-up is reported and is on the entire cohort (e.g., includes 330 ART-experienced)<br>Patients can return to care |
| Masuke 2023 | Sub Saharan Africa | | Urban; Rural | 2016 | July 2013 - June 2014; July 2017 - June 2018 | July 2017 - June 2018 | 9025 | 37 (11.3) | 65.3 | - | No | Includes 11 second line clients |
| Mody 2021 | Sub Saharan Africa | National | Urban; Rural | 2017 | January 1, 2016 - January 1, 2018 | January 1, 2017 - January 1, 2018 | 65673 | 32 (26-39) | 62.2 | 287 (147-466) | No | |
| Mugenyi 2022 | Sub Saharan Africa | | Urban; Rural | 2017 | January 1, 2015 - December 31, 2018 | January 1, 2015 - December 31, 2018 | 20171 | 34.2 (27.6-42.7) | 60.6 | - | No | |
| Mwamuye 2022 | Sub Saharan Africa | | Urban; Rural | 2016 | April 2016 - August 2016; April 2017 - August 2017 | April 2016 - August 2016; April 2017 - August 2017 | 786 | 39.3 (32.5-47.5) | 69 | 355 (172-514) | No | |
| Nimwesiga 2023 | Sub Saharan Africa | | Urban; Rural; Other | 2017 | January 2019 - December 2020 | January 2019 - December 2020 | 80 | 0 | 91.3 | - | No | |
| Ojiambo 2023 | Sub Saharan Africa | 16500 | Urban | 2017 | 2017 - 2020 | 2017 - 2020 | 328 | 36 (IQR = 17) | 68.6 | - | 0 | |
| Ross 2023 | Sub Saharan Africa | | Urban; Rural | 0 | Enrolled in care after Treat-All implementation and prior to January 2019. | Enrolled in care after Treat-All implementation and prior to January 2019. | 29017 | 35 (28-43) | 60.7 | - | No | |
| Singh 2023 | Southeast Asia | | Urban | 0 | July 2016 - December 2017 | July 2016 - December 2017 | 135 | 35.1 (8.97) | 34.1 | - | No | |

| | | | | | | | | | | | |
|---|---|---|---|---|---|---|---|---|---|---|---|
| Tlhajoane 2021 | Sub Saharan Africa | | Rural | 2016 | July 2015 - June 2016; July 2016 - June 2017 | July 2015 - June 2017 | 829 | 38.3 (11.8)<br>37.0 (11.3) | 53.8 | 229.5<br>220 | No | Part of sensitivity analysis<br><br>Assumes the 136 who returned to care and re-entered into cohort after >90 day absence as treatment interruption |
| Zhong 2023 | East Asia | | Urban | 0 | March 1, 2019 - May 31, 2021 | March 1, 2019 - May 31, 2021 | 285 | 37.0 (28.0-54.0) | 10.9 | 84.0 (25.5-145.0) | No | Cannot determine retention at final endpoint, only can determine the 12-month retention |

**Supplemental Table 3.** Definitions of loss to follow-up

| Reference | Definition of LTFU/Retained |
|---|---|
| **Ahmed 2020** | Retention-in-care at 6- and 12-months, which was defined as PLHIV known to be alive and receiving ART at the end of a follow-up periods<br><br>Individuals who did not refill their ART for a period of one or more month after their last refill appointment date and were not yet classified as having died or TO were labeled as LTFU. However, individuals who discontinued treatment but returned to care before 6- and 12-months post-ART initiation were considered as retained at 6-and 12-months, respectively.<br><br>Death - All cause death whose death was recoded on the patient's medical record or ART register |
| **Alhaj 2019** | LTFU: not returned to the clinic two months after the patient is expected to have run out of ART, based on the number of tablets dispensed at the last visit, and is not known to have transferred out, stopped ART, or died.<br>Retention: patient is alive and on ART by the end of the 12-month follow-up period. |
| **Atuhaire 2022** | Failure of a FSW to return to the HIV clinic for ARV drug refill for at least 90 days preceding their last clinic appointment and not classified as transferred out to another clinic for treatment<br><br>Retained in care if they made a clinical visit within 90 days of a scheduled visit. |
| **Avalos 2019** | - |
| **Benzekri 2022** | Retained: alive, retained in care, and receiving ART as determined by patient report and the medical records.<br>LTFU: No contact with the clinic for >6 months and were alive but not retained in care or if they could not be traced. |
| **Byamukama 2022** | - |
| **Cassidy 2023** | 6-Month attrition is defined as a gap of more than 9 months with last visit before gap occurring before month 6 and is only presented for those who initiate ART more than nine months before dataset closure. Those who transferred out<6 months after initiation were excluded<br><br>12-Month attrition is defined as a gap of more than 9 months with last visit before gap occurring before month 12 and is only presented for those who initiate ART more than 15 months before dataset closure. Those who transferred out<12 months after initiation were excluded |
| **Chauke 2020** | LTFU was defined as patients who have not visited the clinic and have not received drugs for more than 90 days after their last clinic visit.<br>Retention was defined as alive and on ART by the end of the 12-month follow-up period. |
| **Dorward 2020** | Attrition was defined as 180 days without a visit |
| **Dumchev 2022** | Treatment Stoppage - 90+ with no engagement after missing a scheduled visit<br><br>Retention - No record of ending treatment 12 months after ART initiation |
| **Eamsakulrat 2022** | Retention in care was defined as having medical records on the day of HIV viral load or CD4 count testing at 12 months after undergoing HIV antibody testing.<br>LTFU was defined as an individual who did not visit the HIV clinic within 3 months after an appointment date and did not have a documented transfer to other care facilities. |

| | |
|---|---|
| **Ntamatungiro 2021** | LTFU was defined as being >60 days late for a scheduled visit |
| **Eshiwani 2018** | - |
| **Harooni 2022** | Defined retained as data indicating the patient visited the center for an ART prescription at 12-15 months after diagnosis |
| **Hirasen 2020** | LTFU was defined as >90 days late for the last scheduled visit with no subsequent visit |
| **Hoang 2022** | LTFU by using the following criteria: (1) being absent greater than 90 days since the last clinic appointment or (2) inadequate information to categorize patients into the group transferred or death |
| **Ibiloye 2018** | Retention: patients link to care at 6 months ART<br>LTFU: lost from the care continuum for more than 2 months since the last appointment |
| **Ibiloye 2021** | LTFU refers to no clinical contact or drug refill from any of the KP-CBART approaches for more than 28 days since the last expected contact<br><br>Attrition - Death or LTFU<br>Retention - Active in care and on ART includes transfers |
| **Izudi 2022** | Retention: Participants with a visit to the HIV clinic at least once within the last four weeks from the date of the last scheduled visit. |
| **Jamieson 2021** | LTFU - >30 days days late for scheduled visit at 18 months (15-21 month window post-ART initiation date window used) |
| **Januraga 2018** | Retention: had at least two outpatient visits at least 90 days apart after ART initiation<br>LTFU: 180 days without any visit to clinic after starting ART |
| **Johansson 2021** | LTUD - >90 days of missing planned clinic visits<br>Retention in care was defined as remaining in care at 12 months after the date of ART initiation, with no registered loss-to-follow-up |
| **Kimanga 2022** | LTFU was defined as individuals missing a clinic visit more than 3 months after their last clinic appointment date. Individuals who transferred care to other facilities were included in the analysis and follow-up time censored at the last clinic visit date before the transfer |
| **Lilian 2020** | LTFU - No drug for 90+ days |
| **Makurumidze 2020** | LTFU - Last recorded clinic visit date or pharmacy pill pick up date was < 180 days before the date of data extraction from the system<br>Attrition - Died, LTFU, Stopped ART |
| **Matare 2020** | LTFU - Not attend last scheduled review visit or pill pick up visit by more than 90 days from date of data collection<br>Attrition - Death LTFU or stopped treatment<br>Retention - Alive and on treatment or transferred out<br>Alive and on Treatment - Known to be on treatment<br>Transferred out - Transfer is documented |
| **Matthews 2020** | LTFU - No contact despite multiple attempts through 13 months after enrollment until device stopped transmitting data or until last scheduled study visit, whichever came first |
| **Mayasi 2022** | LTFU: not presenting to the care center for at least 180 days after the date of a last missing visit (clinical visit, refill of drugs, blood test) without a notification of death or transfer.<br>Retention: alive on ART with no outcome of death or LTFU at the end of data collection |
| **Mshweshwe-Pakela 2020** | Retention: evidence of a care visit (in the same facility where patients had initiated ART) or HIV-related laboratory testing between 91 and 365 days after ART initiation. |
| **Nshimirimana 2022** | LTFU - Failure to report for drug refill within 90 days from last appointment |
| **Onoya 2021** | LTFU - 12 months post diagnosis defined as >90 days late for the last scheduled visit |
| **Opio 2019** | LTFU - A patient who has not visited the health facility HIV clinic in three or more consecutive months at any point in their care since they initiated ART |

| | |
|---|---|
| **Opito 2020** | Retention: active in TASO Tororo care 12 months after initiation |
| **Pillay 2019** | - |
| **Rogers 2021** | Failed to return for drug pick-up after 3 consecutive months following the last expected pharmacy appointment date and were reported as lost to follow-up in the clinic ART registers. |
| **Romo 2022** | LTFU - No record for 6+ months immediately before censoring at 12 months after TB diagnosis, known facility transfer or other documented reason for leaving care with the last recorded data of contact used to determine the timing of the outcome |
| **Ross 2019** | Retention - Having at least 1 health center visit within 5-9 months of enrollment |
| **Seekaew 2019** | - |
| **Seekaew 2021** | Retention - In contact with and refilling ART at ART site 3, 6, and 12 months after initiation which was evaluated by calling participants and checking their ART status in the national system<br>LTFU - Could not be contact via phone and no data registered on the national system 1 month after each specified time point |
| **Ssempijja 2020** | Patients were defined as LTFU if they missed their scheduled ART refill appointments for more than 90 days |
| **Stafford 2019** | LTFU - No documented attendance at the healthcare facility within 90 days from the date of the missed scheduled visit to see a provider or medication pickup |
| **Stockton 2021** | Alive and in Care - On ART for at least some portion of the 2 months prior to 6-month mark starting ART<br>Continuous Engagement - Never more than 14 days late to an appointment through 6 months<br>Currently on ART - Attend an appointment prior to 6-month mark and receiving a supply to last through 6-month mark<br>In care after 6 months - One appointment where ART is provided after 6 months in care |
| **Teshale 2020** | LTFU - Not taking ART refill for 3 months or longer from the last attendance for refill and not yet classified as dead or transferred-out |
| **Bernard Marc 2023** | - |
| **Chagomerana 2023** | Women were considered as LTFU if they could not be reached by telephone or be located in the community on three tracing attempts and did not report to clinic within 3 months of their scheduled date. |
| **Dorward 2023** | LTFU - 90 days late for a visit by the South African ART programme, with date of last visit used as date of loss to follow-up<br>Retention - Not recorded as deceased, LTFU, or transferred |
| **Gemechu 2023** | Any patient who was either not known to have died or had been receiving treatment elsewhere but was absent at the official treatment facility during the regular refilling time where the patient had been registered. After the last missed appointment, the patient had not been seen at the ART clinic for >= 30 days, and healthcare providers were unable to reach them by phone. |
| **Govere 2023** | LTFU - No documented death or transfer and no clinic visit or pharmacy pick-up in the last 90 days<br>Retention - Consistently attending all 1-, 3- and 6-month scheduled clinic visits for treatment collection and clinical management |
| **Hamooya 2023** | Retained: regular attendance at appointments or engagement with the ART clinic at 3, 6, 12, 24, and >= 24 months after ART initiation.<br>LTFU: PWH whose medical records showed no evidence of any form of contact with a health facility for 30 or more days after the scheduled visit day. |
| **Joaquim 2023** | - |
| **Kimanga 2023** | LTFU - Missed scheduled appointments with a 3-month grace period |

| | |
|---|---|
| **Lavoie 2023** | LTFU - No drug pick-up for greater than 28 days after a missed drug pick-up appointment and the patient was not dead, transferred out, or did not return by the end of the study period (e.g, treatment interruption) |
| **MacKellar 2022** | LTFU - Not retained on ART<br>Retained on ART - Not being more than 90 days late for the last antiretroviral refill appointment on the date of abstraction |
| **Masaba 2023** | LTFU - Missed their last appointment by 30 days or more with no documentation of death or transfer out of the study site |
| **Masuke 2023** | LTFU - No clinical contact for 28 days after the last scheduled appointment or expected clinical contact<br>Retention - On ART<br>Opted Out - Stopped ART<br>Transferred ART - Confirmed to have transferred to another health facility |
| **Mody 2021** | Retention - Clinic attendance 9-15 months after enrollment with at least 6-months on ART |
| **Mugenyi 2022** | Retention - Alive and in care<br>LTFU - Not seen more than 3 months from their next appointment date |
| **Mwamuye 2022** | Retention - Alive and on ART<br>Attrition - Discontinued ART, death, and/or LTFU |
| **Nimwesiga 2023** | Retention - Alive and are still receiving health care at study facilities after they were initiated on therapy, includes patients who stopped ART temporarily due to medical/personal reasons<br>LTFU - Missed his or her next clinic or pharmacy refill appointment for at least 1 month and at most 3 consecutive months OR Missed his or her planned clinic or pharmacy refill appointment for more than 3 consecutive months |
| **Ojiambo 2023** | Retention - Consistency in HIV care appointment attendance over time measured as having two and four completed visits at 6 and 12-months respectively<br>Dropped - Missing their next appointment for 1-2 months<br>Lost - Missing next appointment for 3+ months in a row without returning<br>LTFU - Died, Dropped, or List |
| **Ross 2023** | LTFU - No contact with the health center for more than 180 days among patients not known to have died or transferred |
| **Singh 2023** | - |
| **Tlhajoane 2021** | - |
| **Zhong 2023** | - |

**- Denotes that a definition was not provided in the text**
